# Differential vulnerability of the cerebellum in healthy ageing and Alzheimer’s disease

**DOI:** 10.1101/2020.02.04.20019380

**Authors:** Helena M. Gellersen, Xavier Guell, Saber Sami

## Abstract

Recent findings challenge the prior notion that the cerebellum remains unaffected by Alzheimer’s disease (AD). Yet, it is unclear whether AD exacerbates age-related cerebellar grey matter decline or engages distinct structural and functional territories. We performed a meta-analysis of cerebellar grey matter loss in normal ageing and AD. We mapped voxels with structural decline onto established brain networks, functional parcellations, and along gradients that govern the functional organisation of the cerebellum. Importantly, these gradients track continuous changes in cerebellar specialisation providing a more nuanced measure of the functional profile of regions vulnerable to ageing and AD. Gradient 1 progresses from motor to cognitive territories; Gradient 2 isolates attentional processing; Gradient 3 captures lateralisation differences in cognitive functions. We identified bilateral and right-lateralised posterior cerebellar atrophy in ageing and AD, respectively. Age- and AD-related structural decline only showed partial spatial overlap in right lobule VI/Crus I. Despite the seemingly distinct patterns of AD- and age-related atrophy, the functional profiles of these regions were similar. Both participate in the same macroscale networks (default mode, frontoparietal, attention), support executive functions and language processing, and did not exhibit a difference in relative positions along Gradients 1 or 2. However, Gradient 3 values were significantly different in ageing vs. AD, suggesting that the roles of left and right atrophied cerebellar regions exhibit subtle functional differences despite their membership in similar macroscale networks. These findings provide an unprecedented characterisation of structural and functional differences and similarities in cerebellar grey matter loss between normal ageing and AD.

## 1. Background

Despite the earlier notion that the cerebellum is spared in Alzheimer’s disease (AD), recent studies have started to elucidate detrimental effects of AD on cerebellar structure, which in some cases correlates with clinical disease ratings (Gellersen et al., 2017; Guo et al., 2016a; Jacobs et al., 2018; Serra et al., 2017; Toniolo et al., 2018). This is in line with the current understanding that the cerebellum is involved in a range of cognitive, motor, and affective functions by virtue of segregated cerebro-cerebellar loops (Kelly & Strick, 2003; Schmahmann, Guell, Stoodley, & Halko, 2019). Cerebellar grey matter loss does not only occur in AD, but the healthy ageing process is also associated with changes in cerebellar structure, which are comparable in magnitude to those observed in the prefrontal cortex and hippocampus (Bernard & Seidler, 2014; Jernigan et al., 2001; Raz & Rodrigue, 2006). These marked alterations highlight the importance of the cerebellum for our understanding of both cognitive ageing and Alzheimer’s disease processes.

However, it is unclear if these AD-related changes exacerbate existing negative effects of ageing on the cerebellum or whether they also target regions less vulnerable to normal ageing processes. Moreover, a direct comparison of the functional characterisation of regions affected by ageing and AD is also still lacking. Here we combine a coordinate-based meta-analytic approach to identify the most consistent patterns of grey matter loss across studies with multiple complimentary functional mappings to advance our understanding of cerebellar vulnerability to normal ageing and Alzheimer’s disease (Buckner, Krienen, Castellanos, Diaz, & Yeo, 2011; Eickhoff, Bzdok, Laird, Kurth, & Fox, 2012; Guell et al., 2019; Guell, Schmahmann, Gabrieli, & Ghosh, 2018; King, Hernandez-Castillo, Poldrack, Ivry, & Diedrichsen, 2019).

The importance of the cerebellum for multiple cognitive, affective and motor domains is increasingly being recognised (Schmahmann et al., 2019), as is its role in age- and AD-related functional changes (Bernard et al., 2020; Bernard & Seidler, 2014; Gellersen et al., 2017; Jacobs et al., 2018). Despite the striking uniformity of cerebellar cellular organisation, the anterior-posterior functional continuum from motor to cognitive regions and the unique cortico-limbic connectivity patterns of each cerebellar lobule suggest non-uniformity of age and pathological effects across this brain region (Bernard, Leopold, Calhoun, & Mittal, 2015; Buckner et al., 2011; Koppelmans et al., 2017; Ramanoël et al., 2018). Indeed, effects of AD are not uniform across cerebellar lobules. In a previous meta-analysis, consistent patterns of AD-related cerebellar atrophy across studies were found in posterior cerebellar lobules (Gellersen et al., 2017), although since then other authors demonstrated that grey matter loss may also occur in anterior lobules of the cerebellum in AD (Toniolo et al., 2018).

An important question concerns the interpretation of these structural findings with respect to potential consequences for function. There are now multiple maps of cerebellar functional organisation derived from task-based fMRI to identify discrete functional boundaries (King et al., 2019), or based on resting-state intrinsic connectivity which define different brain networks (Buckner et al., 2011) or explain connectivity patterns in terms of a region’s relative position along major cerebellar functional gradients, respectively (Guell et al., 2018). Three major gradients have been shown to govern the functional architecture of the cerebellum: the first tracks the transition between cognitive to motor regions, the second defines the extent to which a region is engaged in a task-positive processing thereby isolating attentional components, and the third captures laterality asymmetries in the left compared to the right cerebellar hemisphere that are specific to non-motor processes. As such, the third gradient suggests that cognitive processing in left and right hemispheres may be qualitatively different, a notion that may not be visible when only using the established network-specific mappings of the cerebellum (Buckner et al., 2011). The unique feature of the gradient-based approach is that each cerebellar voxel is characterised based on its position along the continuum between the two extremes of each functional gradient. As a result, voxels do not have to be categorised into one of the functional networks or parcels (Buckner et al., 2011; King et al., 2019), but can take on any position along the functional spectrum allowing for a comparison of the relative functional preferences of regions vulnerable to AD and normal ageing.

All of these approaches provide complimentary information about cerebellar functional organisation: the network-based approach puts cerebellar regions into context with the functional characterisation of the neocortex (Buckner et al., 2011); the discrete parcellation allows for a direct mapping of specific cognitive, motor and affective processes onto cerebellar anatomical subregions (King et al., 2019); and the gradient-based approach presents a picture of gradual changes in functional preferences across major functional axes, refraining from defining discrete boundaries and allowing each region to be characterised by multiple features (Guell et al., 2018). These gradients also provide information regarding the relationship between different functional territories of the cerebellar cortex, an important piece of information which is absent in discrete maps of cerebellar functional neuroanatomy. The use of all three methods for the interpretation of functional consequences of grey matter atrophy is therefore synergistic.

Yet, in most studies, the interpretation of implications of age- or AD-dependent structural decline for cerebellar function is based on the location of the identified cluster within a given lobule (Bernard et al., 2015; Bernard & Seidler, 2013; Buhrmann et al., 2020). While valuable, such an approach may miss further subtler functional differences within a region or continuities between regions that only become apparent on non-discrete mappings. Moreover, functional territories do not neatly map onto macroscale anatomical boundaries (King et al., 2019).

Here, we used the discrete and network-based maps in combination with the gradient-based approach which allowed us to place our meta-analytic structural imaging findings on multiple functional maps and to compare those derived from ageing with those from AD. The use of gradients can highlight even subtle functional differences between neuroanatomical regions vulnerable to AD and normal ageing (Guell et al., 2018), which can be put into context with discrete mappings that have typically been used by most prior studies. These investigations will further our understanding of the role that grey matter decline in the cerebellum may play in functional deficits and will demonstrate whether these patterns of age-related grey matter loss are distinct from those in Alzheimer’s disease.

## 2. Methods

### 2.1 Literature Search

We carried out a systematic literature search on Pubmed and PsycInfo using search terms related to ageing and structural neuroimaging in the title and/or abstracts of peer-reviewed records that were published until October 1^st^ 2019. Inclusion criteria required 1) an analysis that compared grey matter volume in healthy young (< 40 years) and older participants (60+ years), and 2) availability of coordinates of regions with grey matter decline (either from a whole-brain analysis or from a region of interest analysis that included the cerebellum). Studies were excluded if 1) participants had psychiatric or neurological conditions, 2) the cerebellum was excluded from the analysis, 3) the study used confounding factors besides covariates sex, age and education in the analysis, and 4) the study reanalysed previously published data already included in our meta-analysis. We extracted sample size, participant demographics, coordinates of cerebellar grey matter loss and information regarding the statistical analysis from each included study. All coordinates reported in Talairach space were converted to MNI space using the icbm2tal transform employed by the conversion tool provided by GingerALE (brainmap.org).

*N*=18 studies were identified for inclusion in the meta-analysis. Input for the analysis were coordinates of age-related decreased cerebellar grey matter volume or density and the sample size of each study. We also conducted an updated analysis of cerebellar grey matter decline in late-onset AD in *n*=13 studies based on our previous research (Gellersen et al., 2017) and studies published since then (Ahmed et al., 2019; Toniolo et al., 2018). The details of the literature search for the AD meta-analysis are shown in Gellersen et al. (2017). Studies with early-onset AD and atypical forms of AD were excluded from the meta-analysis to reduce heterogeneity in patient samples.

For details on search terms and the PRISMA flowchart for study selection see Appendix A. For a list of participants, imaging parameters, cerebellar coordinates, cognitive deficits (age- and AD-related) and associations between cerebellar grey matter and cognitive performance see Appendix B (Table B1 for healthy ageing and Table B2 for AD).

### 2.2 Meta-Analysis

The coordinate-based random-effects meta-analyses on age and AD were carried out using anatomical likelihood estimation (ALE) using version 2.3.6 of GingerALE (http://brainmap.org) (Eickhoff et al., 2012, 2009; Turkeltaub, Eden, Jones, & Zeffiro, 2002). In the coordinate-based meta-analysis, every coordinate, referred to as a “focus”, is treated as the centre of a Gaussian probability distribution with a full-width half-maximum based on the sample size of the respective experiment. For each study, a modelled activation (MA) map of grey matter differences between age groups was formed by finding the maximum across the Gaussian probability distributions of all foci. The final ALE image that represents the results of the meta-analysis is created from the union of the MA maps of all studies. To determine the null distribution of the ALE statistic, values from the MA maps are sorted to create histograms. Histograms are then divided by the number of voxels in a map to generate probabilities for each value of the MA map. Based on these, *p*-value images are created which can then be used for thresholding.

For the current analysis, a cluster-level thresholding method was chosen. This method simulates a random data sets with the same characteristics of the original data set used to create the original ALE image, namely the total number of foci, number of foci groups, and sample sizes. A probability threshold is then set by selecting a minimum number of voxels per cluster in a way that only a certain amount of the clusters in the simulated data sets exceed this minimum size. In the case of this analysis, we chose a *p*<.05 for cluster-level family-wise error correction (i.e. the cluster-level inference option on GingerALE), which provides the best compromise between sensitivity and specificity (Eickhoff et al., 2016). The cluster-forming threshold was chosen to be *p*<.001 (uncorrected), which is one of the two thresholds recommended by GingerALE (Laird et al., 2005). To demonstrate the convergence between these analyses, we created a conjunction image in GingerALE based on the voxelwise minimum between the two thresholded ALE images derived from the AD and the ageing analyses (Eickhoff et al., 2012, 2011).

It is important to note that a region of interest (ROI) based meta-analysis does not provide information about the likelihood or prevalence of cerebellar atrophy in older adults and AD but rather characterises the pattern of lobular grey matter loss in cases where cerebellar atrophy is indeed present.

### 2.3 Visualisation of meta-analytic results

We used the SUIT toolbox to create multiple cerebellar maps, which are created by unfolding and flattening the cerebellar surface to obtain a 2D representation (Diedrichsen, Balsters, Flavell, Cussans, & Ramnani, 2009; Diedrichsen et al., 2011; Diedrichsen & Zotow, 2015). Note that the flatmap is not a true surface reconstruction and is based on volumetric MRI data (Diedrichsen & Zotow, 2015).

To put our structural findings into context, used three maps of the functional organisation of the cerebellum that have been derived on the basis of cerebellar functional activation and cortico-cerebellar connectivity patterns: 1) the Buckner networks based on intrinsic connectivity derived from resting-state fMRI showing the membership of different cerebellar subregions to one of the established functional connectivity networks of the brain (Buckner et al., 2011), 2) the King et al. (2019) map based on a multi-domain cognitive task battery, and 3) a gradient-based, continuous map of the functional preferences of different cerebellar regions (Guell et al., 2018), which can be used to quantify relative differences in the functional characteristics of cerebellar voxels (see section 2.4).

### 2.4 Using gradients to quantify relative differences in functional preferences of regions with grey matter loss in ageing and Alzheimer’s disease

To further add to the topographical interpretation of our meta-analytic findings, we used a recently developed tool called LittleBrain (Guell et al., 2019; https://xaviergp.github.io/littlebrain/). LittleBrain maps the voxels identified in our structural analysis onto gradients that capture low-dimensional properties of cerebellar functional organization that has previously been described using a non-linear dimensionality reduction method called diffusion map embedding on Human Connectome Project functional resting-state data (Guell et al., 2018; Margulies et al., 2016). The method identified the functional relationships of intrinsically connected cerebellar voxels, which may not be directly connected through cerebellar neocortical association fibres but likely participate in similar functional cerebro-cerebellar loops. Diffusion map embedding defined components that account for as much of the variability in the data as possible, similar to linear methods such as principal component analysis, with the crucial distinction that voxels were not assigned to separate networks. This mapping method therefore overcomes the limitation of being constrained by discrete boundaries established through structural parcellation and can identify the progressive hierarchical functional organisation across cerebellar regions as has previously been achieved for the neocortex (Margulies et al., 2016). The method allowed for overlap between functional gradients such that each voxel has a position in each gradient (Guell et al., 2018). Note that voxels that are close in the gradient dimensions exhibit similar functional connectivity patterns.

The gradient-based map demonstrated that the functional macroscale organisation of the cerebellum can be characterised as following along two principal gradients. Cerebellar Gradient 1 represents the principal functional axis of the cerebellum, including motor representations on one extreme (lobules IV-VI and VIII) and transmodal cognitive processing such as cerebellar default mode network regions on the other (Crus I/II and lobule IX). Gradient 2 isolates attentional processes moving along a task-focused to task-unfocused functional axis (Guell et al., 2018). One end of this gradient includes anterior regions Crus I/II involved in the frontoparietal and attention networks, while the other end includes motor (lobules I-IV) and DMN regions (posterior Crus I/II). We also chose to include Gradient 3, which captures more subtle aspects of the functional profile of cerebellar regions. Specifically, it demonstrates differences in lateralisation of non-motor functions. This is relevant given that our findings for AD and ageing exhibit differences in hemispheric lateralisation.

After identifying regions of grey matter loss, we created plots of the position that each of the voxels with grey matter loss in aging, AD and the Aging-AD conjunction held along the three functional gradients. Furthermore, each voxel was colour-coded according to its participation in the cerebro-cerebellar resting-state networks identified in Buckner et al. (2011).

Gradient values were extracted for further statistical analyses to determine whether the distribution along each gradient for the aging and AD effects differed from one another. Due to non-normality of the true gradient value distributions (Shapiro-Wilk tests: *W*>.8, *p*<.001), differences between the AD and ageing gradient values were tested using permutation testing of differences in rank sums. Differences between the AD and ageing gradient values were investigated using permutation testing. Gradient values were assigned to the AD and ageing group randomly and the rank sums between the real observed and random differences between the two groups were tallied. The reported *p*-value for the permutation test represents the probability of getting the observed differences in gradient values between the true AD- and true age-related voxels from random group assignments of each voxel in the permutation procedure.

We also computed the non-parametric effect sizes Cohen’s *U*_*1*_ (Cohen, 1988) using the Matlab toolbox “Measures of Effect Size” (Hentschke & Stüttgen, 2011): Cohen’s *U*_*1*_ can be interpreted as the proportion of scores across both groups within the areas of no overlap, such that completely separate distributions result in *U*_*1*_=1 (large effect) and completely overlapping distributions have *U*_*1*_=0 (null effect) (Cohen, 1988; Hentschke & Stüttgen, 2011).

The distributions of gradient values were visualised using violin plots in ggplot2 in RStudio version 1.0.153 (Wickham, 2016).

### 2.5 Robustness tests: jackknife procedure

The robustness of our results for the ageing and AD analyses, respectively, to the removal of studies was tested using a jackknife procedure, in which the meta-analysis was carried out without a given study. As a result, 18 one-study removed analyses were carried out for the ageing dataset and 13 such analyses were carried out for the AD dataset, in which we conducted the ALE meta-analysis without a given study. We also tested whether the difference in the distribution of AD gradient values and the jackknife analysis gradient values remained robust to the removal of any one study.

## 3. Results

### 3.1 Healthy ageing meta-analysis

The ageing analysis involved 18 studies with 64 coordinates of cerebellar grey matter atrophy across 2441 subjects (Appendix B, Table B1 for details of included studies). Five clusters of reduced grey matter volume were identified (Figure 1a, Table 1; all local peaks *p*<.020). Cluster 1 was situated in right Crus II. Cluster 2 encompassed parts of right Crus I and lobule VI. Cluster 3 was in left Crus II. The fourth cluster of grey matter volume reduction was in vermal lobule VI, encroaching upon left hemispheric lobule VI. The fifth cluster was located in lateral left posterior hemisphere with peaks in Crus II and Crus I, with some extension into lobules VIIb and VIII.

**Table 1.**
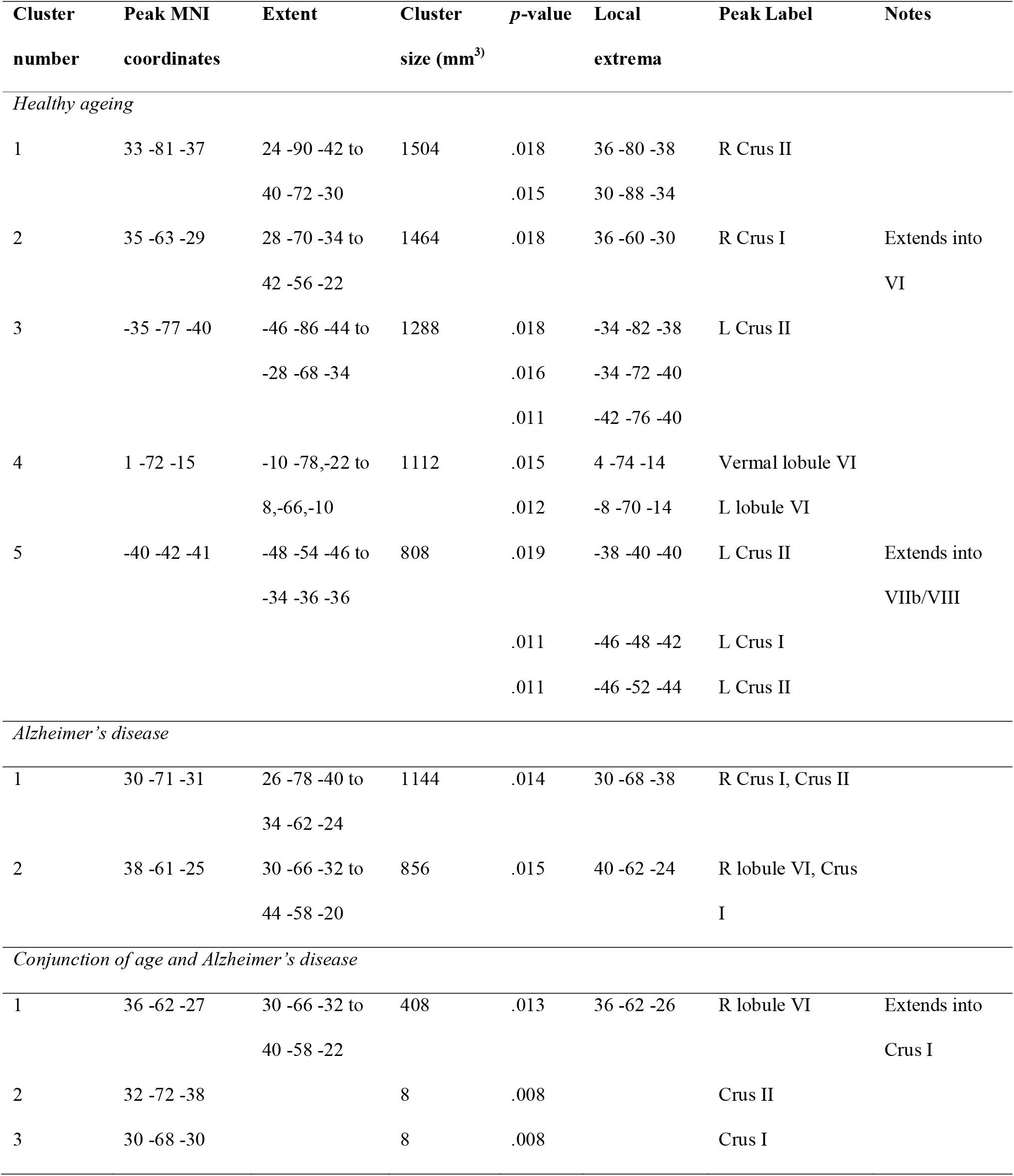
Results of the anatomical likelihood estimation meta-analyses for healthy ageing (*n*=18 experiments with a total of 2441 subjects and *k*=64 foci of cerebellar grey matter decline), Alzheimer’s disease (*n*=13 experiments with a total of 529 subjects and *k*=35 foci of cerebellar decline), and the conjunction which identified the spatial overlap between effects of Alzheimer’s disease and age on cerebellar structure. Here we report the Montreal Neurological Institute coordinates returned by GingerALE. Labels are according to the spatially unbiased atlas template of the cerebellum and brainstem (SUIT).

**Figure 1.**
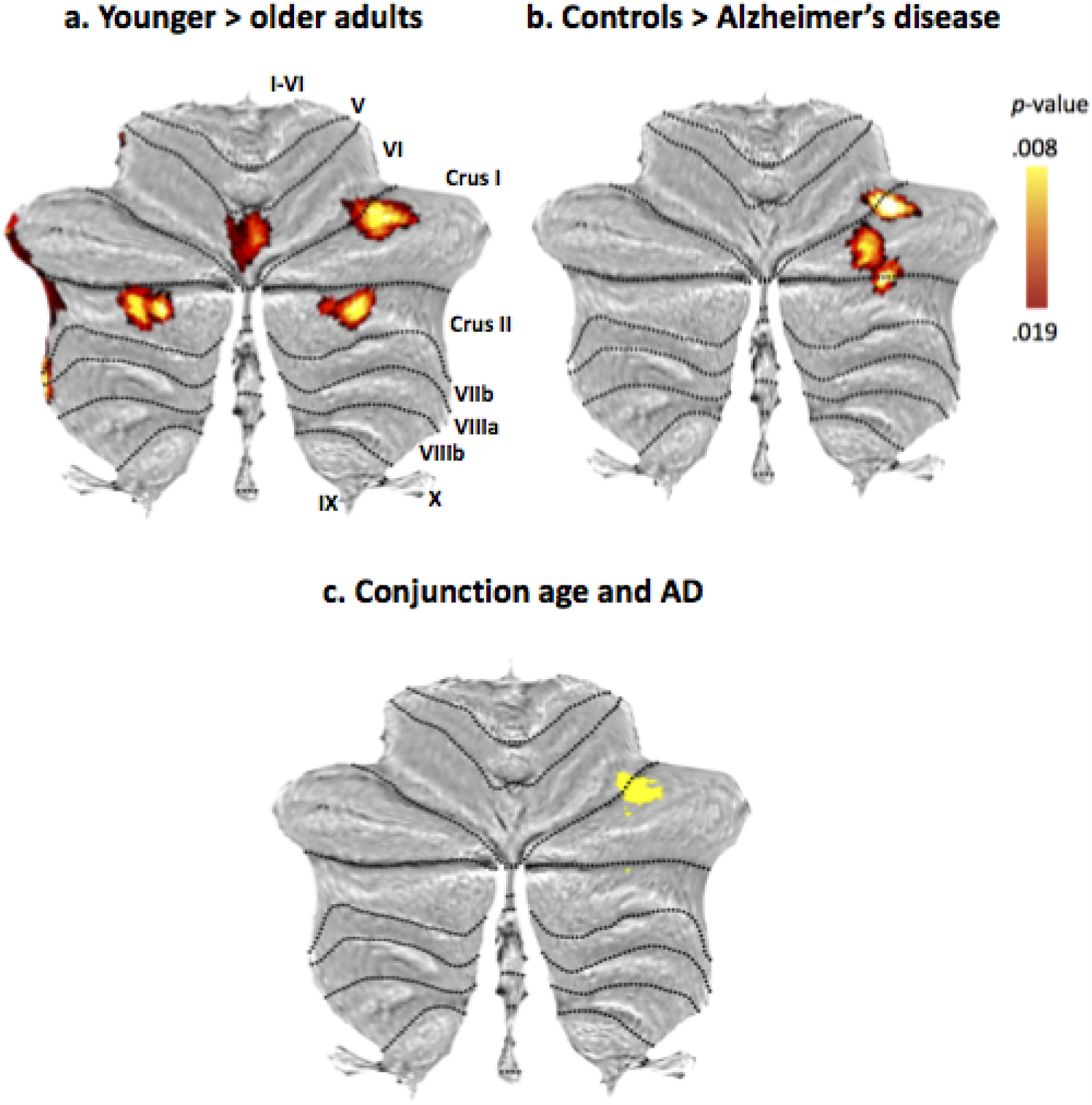
(a) Cerebellar grey matter loss in across 18 healthy ageing studies. (b) Cerebellar grey matter reductions in Alzheimer’s disease across 13 studies. (c) Conjunction of effects of healthy ageing and Alzheimer’s disease on grey matter volume. Note that these results are based on the voxelwise minimum value from the two input analyses on ageing and AD.

### 3.2 Alzheimer’s disease meta-analysis

The final dataset for the AD analysis comprised *n*=13 studies in 529 patients and controls with *k*=35 foci of cerebellar grey matter loss relative to controls. Study characteristics for the included studies can be found in Appendix B (Table B2). The meta-analysis revealed two adjacent AD-related cluster of grey matter loss in the right posterior hemisphere (Figure 1b; Table 1; all *p*<.020). Cluster 1 included regions of right Crus I and II. Cluster 2 comprised regions of lobule VI and Crus I. The inclusion of the additional studies in this analysis compared to our previous results increased the extent but did not have a substantial effect on the location of the peak coordinates (Gellersen et al., 2017).

### 3.3 Conjunction between ageing and Alzheimer’s disease

The conjunction image generated by GingerALE to compare the AD and ageing results demonstrated that there was one larger region of overlap between these two groups (*p*=.013; Figure 1c; Table 1; see also two minor regions of overlap). This was a cluster in right Crus I/lobule VI that had emerged in both analyses.

### 3.4 Placing our meta-analytic structural results in context with functional gradients and networks of the cerebellum

For plots comparing distributions of gradient values for each single gradient between age, AD and the AD-age overlap see Figure 2. Figure 3 shows the distribution of gradient values colour-coded by functional brain network and flatmaps of gradient distributions with AD- and age-related atrophy clusters superimposed.

**Figure 2.**
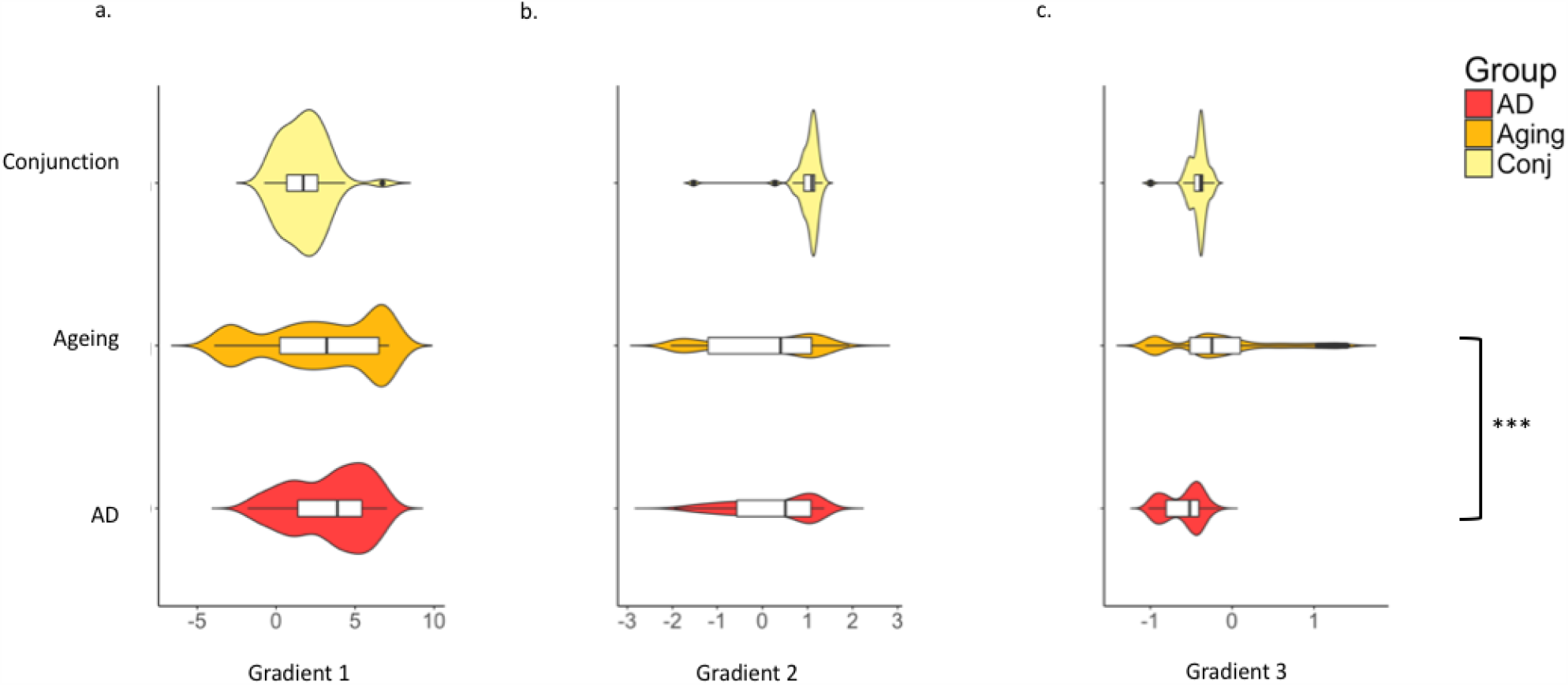
Violin plots showing the distribution of gradient values by analysis group. Permutations testing for differences in the rank sum of gradient values between healthy ageing and AD found no difference in Gradients 1 (a) and 2 (b) values. A significant difference in mean values was found for Gradient 3 (c) values. **\*\*\* *p*<**.**001**

**Figure 3.**
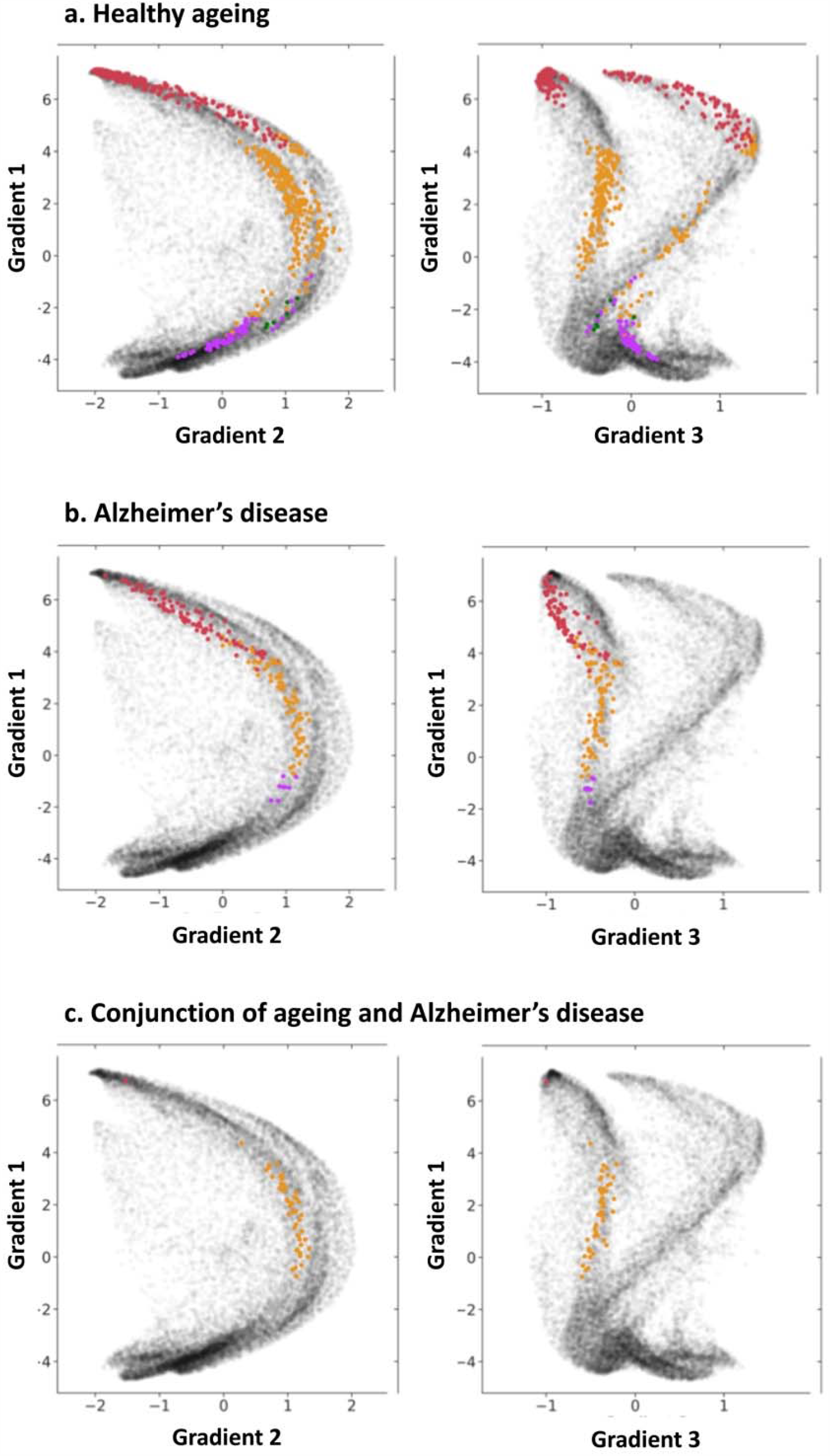
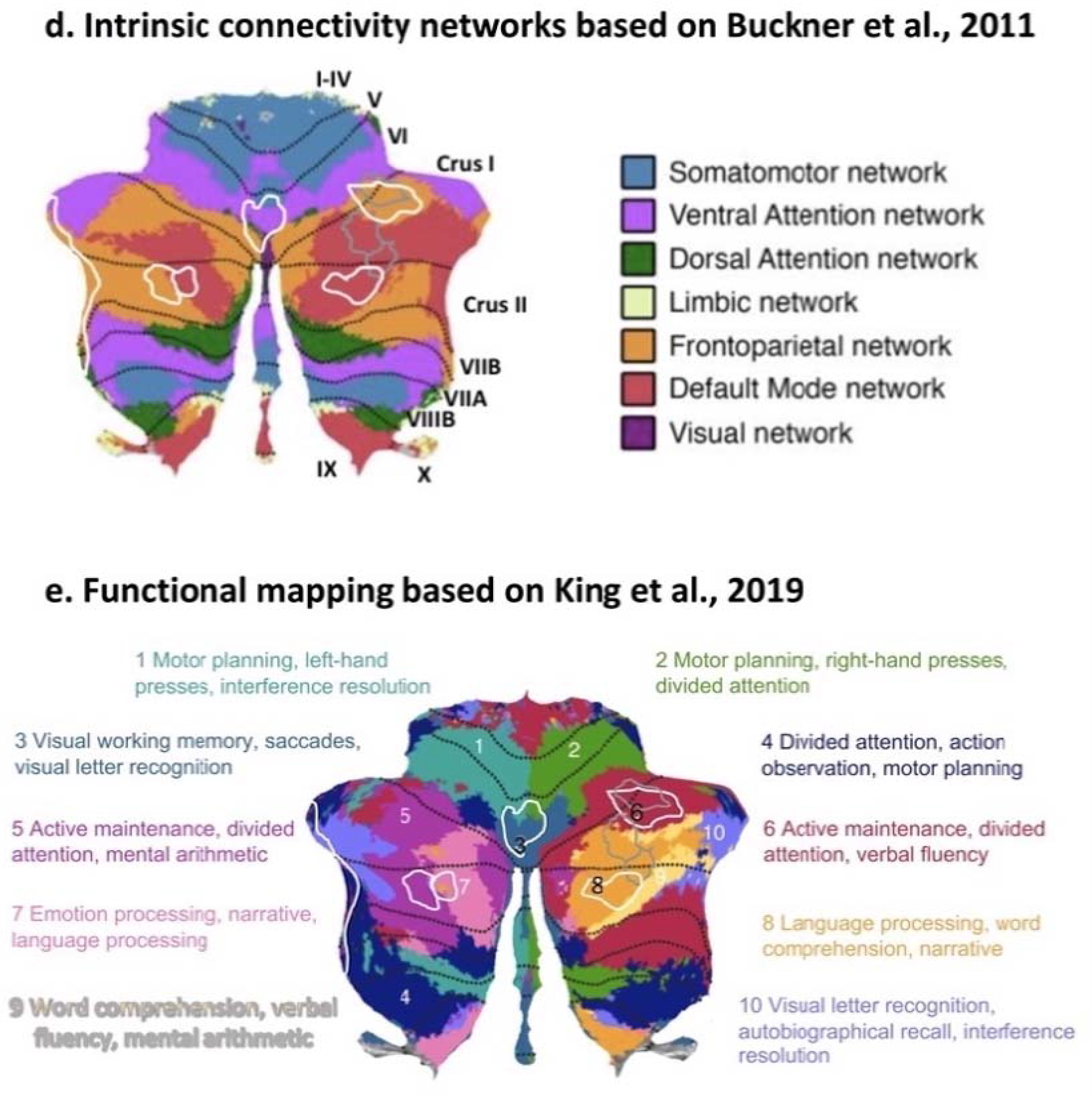
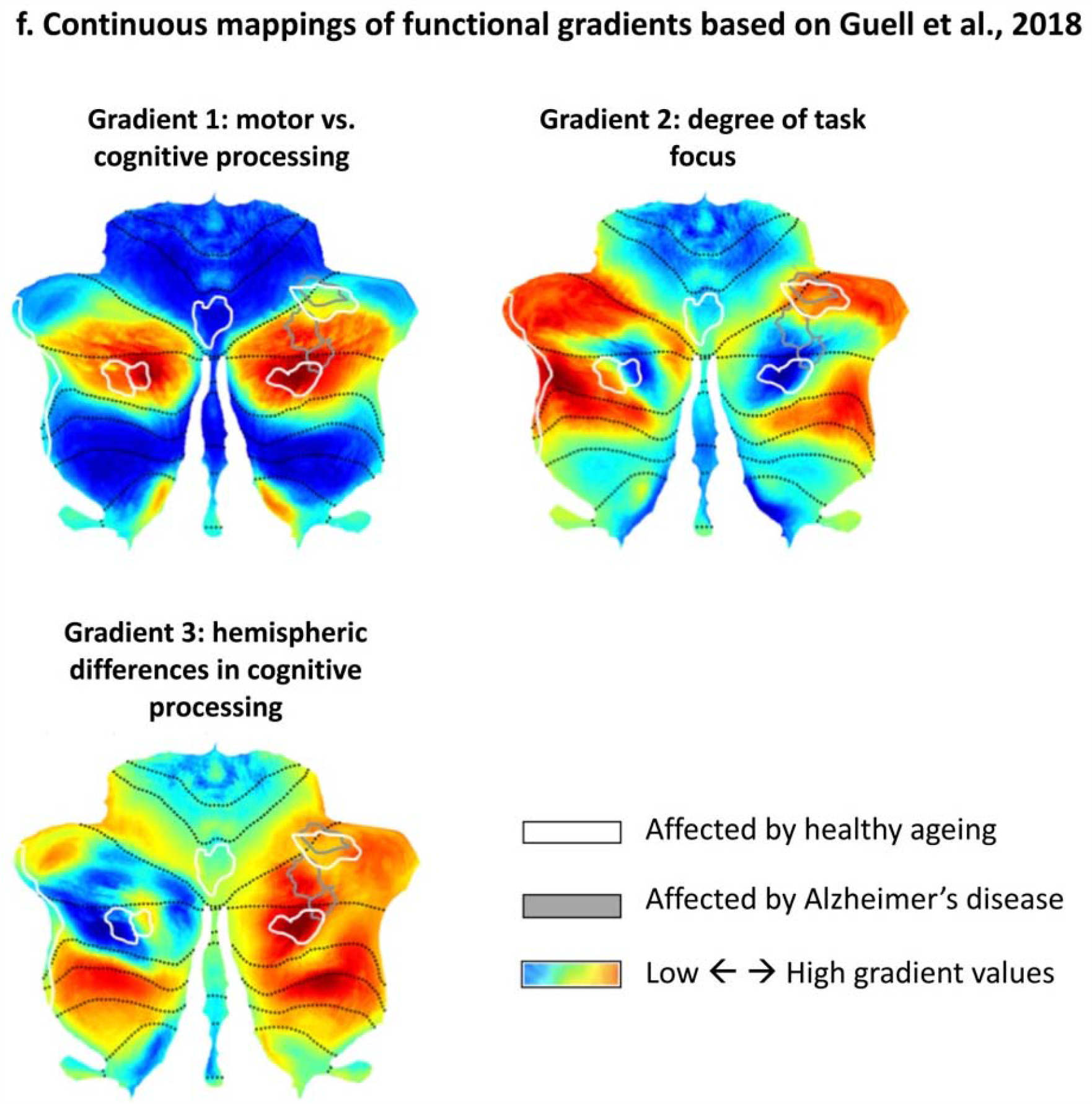
The distribution of the voxels of age-related grey matter loss according to the three main functional gradients of the cerebellum for (a) healthy ageing, (b) Alzheimer’s disease and (c) the conjunction of ageing and AD. Colour codes represent the intrinsic resting-state networks identified in Buckner et al. (2011) which are shown on a cerebellar flatmap in (d) (Diedrichsen and Zotow, 2015). (e) Task-based functional map of the cerebellum from King et al. (2019). (f) Gradient-based maps based on Guell et al. (2018). Gradient 1 represents the transition from motor tasks (lower values) to task-unfocused cognitive processing such as in the default mode network (higher values). Gradient 2 isolates attentional processing moving from task-unfocused (low values) to task-focused (high values) processes. Gradient 3 shows differences in lateralisation of non-motor processes. The regions with grey matter loss identified in our meta-analysis are plotted on top of each flatmap for healthy ageing (white outline) and Alzheimer’s disease (grey outline).

A comparison of the clusters of age-related grey matter reduction from our meta-analysis with the network connectivity patterns derived from Buckner and colleagues (Buckner et al., 2011) showed that the majority of age-related structural decline occurred in regions belonging to the frontoparietal and default mode networks (DMN), with some involvement of ventral and dorsal attention networks (Figure 3). The voxels in the age-related grey matter loss clusters were involved in cognitive processing (tendency towards higher Gradient 1 values, *M*=2.833, *SD*=3.630) and encompassed both task-focused and task-unfocused domains (Gradient 2 mean value close to zero, *M*=.055, *SD*=1.193). Gradient 3 values for ageing included both minimum and maximum values, reflecting the distribution of age effects across the two hemispheres in cognitive territories of the cerebellar cortex (*M*=-.144, *SD*=.652). According to the overlap of age-related grey matter loss and the King et al. (2019) map, age-affected regions participate in working memory, attention, visual, affective and language processing.

The regions affected by AD also participate in DMN and the frontoparietal network. Higher Gradient 1 values highlight the involvement of cerebellar cognitive regions in this disease (*M*=3.381, *SD*=2.427), while motor regions are spared. The range of Gradient 2 values implicates both task-focused and unfocused processing regions in AD (*M*=.220 *SD*=.929). Gradient 3 values for AD were negative, reflecting the right lateralisation of grey matter decline (*M*=-.604, *SD*=.228). According to the overlap of age-related grey matter loss and the King et al. (2019) map, regions with AD-related grey matter atrophy participate in working memory, attention and language processing.

The ageing-AD conjunction voxels were located in regions that exhibit greater involvement in task-focused cognitive processes (high Gradient 2 values; *M=*.995, *SD*=1.193; Figure 3c). While grey matter loss in both AD and ageing involved DMN regions, all conjunction voxels overlapped with areas previously identified as belonging to the frontoparietal network (Buckner et al., 2011), in line with their role in cognitive control processes, such as divided attention and verbal fluency (King et al., 2019).

The distributions of Gradient 1 and 2 values in healthy ageing and AD were highly overlapping (Gradient 1: *Z*=.320, *p*=.748; Cohen’s *U*_*1*_=.203; Gradient 2: *Z*=1.105, *p*=.272; Cohen’s *U*_*1*_=.084; Figures 2, 3). In contrast, regional grey matter loss due to healthy ageing was associated with higher median Gradient 3 values compared to AD-related structural decline (*Z*=-10.549, *p*<.001; Cohen’s *U*_*1*_=.377), reflecting the involvement of only the right (AD) as opposed to left and right (ageing) cerebellar hemispheres in each group. This finding could not be identified only based on macroscale brain network membership (DMN, FPN; Figure 3d), but was reflected in the discrete functional mapping which suggested differences in the extent to which left and right hemispheres were involved in affective and language processing, respectively (Figure 3e).

These findings regarding AD- and age-related cerebellar atrophy in regions of the DMN and FPN dovetail with cognitive deficits the included studies reported for older adults and AD patients, respectively. Across a variety of cognitive tasks, both healthy ageing and AD were associated with impairment in episodic and working memory as well as executive functioning (Appendix B, Table B1 for healthy ageing and Table B2 for AD). AD patients also exhibited deficits in verbal fluency. The studies on healthy ageing included here did not conduct further analyses to establish behavioural correlates of specific cerebellar regional grey matter decline. Among the few studies relating grey matter to clinical measures in AD, no correlation between MMSE scores and cerebellar atrophy was found (Colloby, O’Brien, & Taylor, 2014; Farrow et al., 2007; Möller et al., 2013; Toniolo et al., 2018), but performance on figure copy tests was associated with volume of both anterior and posterior lobes (Toniolo et al., 2018).

### 3.5 Robustness tests: jackknife analyses

The detailed results of each jackknife procedure are reported in Appendix D. For both the AD and ageing jackknife analyses there was no case in which removing one study from the analysis changed the label of the peak voxel of a cluster (Appendix D, Tables D1 and D2). Only slight shifts in peak voxel locations were observed. The jackknife procedure showed that healthy ageing clusters 1 and 2 in right Crus I/II were robust to the removal of any one study. Clusters 3 and 4 survived removal of all except one study each and remained stable in all other instances, indicating robustness in 94% of analyses. Cluster 5 in anterior Crus I/II remained stable in 83% of all analyses but did not survive correction for multiple comparisons upon removal of studies with a larger number of foci. For AD, there were two instances in which the removal of a study affected the survival of one of the two clusters, but never both simultaneously. As a result, there was not a single jackknife analysis in which no AD-related atrophy occurred in the cerebellum and each cluster was stable in 85% of analyses. In five jackknife analyses, Clusters 1 and 2 merged into one larger cluster without resulting in substantial shifts in the locations of cluster peaks.

On average across all jackknife analyses, Gradient 1 and 2 values were not significantly different between healthy ageing and AD. The significant difference in Gradient 3 between ageing and AD remained in all leave-one-out analyses. In all jackknife analyses, age- and AD-related regions were part of the DMN and FPN networks and were involved in cognitive control and language processing, demonstrating the stability of the functional mappings of regions with AD- and age-dependent grey matter loss.

## 4. Discussion

### 4.1 General Discussion

We aimed to determine whether Alzheimer’s disease and normal ageing result in dissociable patterns of cerebellar grey matter loss and whether the functional profile of the affected regions could be differentiated. To answer these questions, we employed anatomical likelihood estimation (ALE) meta-analysis to identify consistent patterns of cerebellar grey matter atrophy across 18 studies of healthy ageing. We further compared age-related decline to that observed in 13 studies of Alzheimer’s disease. Age and AD exhibited grey matter decline in posterior cerebellum in Crus I/II and lobule VI, albeit in largely non-overlapping subregions of these lobules. The most striking finding was the strict right lateralisation of grey matter loss in AD compared to bilateral decline in healthy ageing.

We used three complimentary mappings of the cerebellum to characterise the functional profile of the regions with grey matter loss in ageing and AD: the known intrinsic brain networks based on resting state fMRI (Buckner et al., 2011), the task-based mapping with discrete functional boundaries (King et al., 2019), and a multi-dimensional continuous map reflecting the three major gradients that govern the functional organisation of the cerebellum (cognitive to motor, degree of task focus, and qualitative differences in left and right homologues of posterior cognitive cerebellum; Guell, Schmahmann, Gabrieli, et al., 2018). Using the gradient-based approach in combination with structural findings allowed a more nuanced characterisation of vulnerable cerebellar regions, over and above discrete or categorical descriptions of their function. It also allowed for a statistical quantification of differences in functional preferences between AD- and age-affected regions.

Despite only a small spatial overlap of regions with age- and AD-related cerebellar grey matter loss, both exhibit a strong bias towards cognitive as opposed to motor processing (Gradient 1) and involvement across the task-focused to unfocused continuum (Gradient 2; Guell, Schmahmann, Gabrieli, et al., 2018). This was also in line with the role of these regions in default mode and frontoparietal networks (Buckner et al., 2011), as well as many shared functional characteristics suggesting their involvement in working memory, attention, and language processing (King et al., 2019).

Notwithstanding these functional similarities, regions affected by normal ageing and AD also showed some differences in their functional characterisation. Specifically, they held different positions along the third gradient. Gradient 3 highlights differences that exist in cognitive processing in the two hemispheres but are not present in motor domains. Note that the interpretation of Gradient 3 values does not simply pertain to spatial localisation of a cluster to the left or right hemisphere. In fact, Gradient 3 reflects higher functional lateralisation of cognitive functions as opposed to primary motor functions. The same regions in left and right homologues of the motor cerebellum (lobules IV-VIII) would be described with the same gradient values. However, left and right hemispheric areas of nonmotor processing (lobules Crus I/II, VIIb, IX) lie on two opposing ends of the spectrum (Guell et al., 2018).

These findings suggest that regions with grey matter loss in ageing and AD do not only reflect differences in spatial localisation to the left and right hemispheres, but also qualitative differences in the functions carried out by the left and right homologues of posterior cerebellar cortex. They point to a dissociation between the functional connectivity fingerprint of the two homologues of posterior cerebellar lobules and highlight their differential vulnerability to AD and age, respectively. In contrast, the intrinsic functional connectivity networks based on resting-state fMRI grouped both regions into the same networks (frontoparietal and default mode; Buckner et al., 2011). The gradient findings provide a complementary statistical confirmation of the picture obtained from the King et al. (2019) map. This map also suggests laterality differences where the right posterior cerebellar hemisphere holds a more prominent role in verbal processing (especially Crus I/II), while only the left region seems to be involved in emotional processes.

### 4.2 Prior evidence of macroscale cerebellar changes in healthy ageing and AD the implications for behaviour

Dominant views on the role of cerebellar function in cognitive, motor and affective processes postulate that the striking uniformity of cerebellar micro-circuitry suggests that a single computational mechanism, the “universal cerebellar transform”, operates on inputs from extra-cerebellar structures involved across behavioural domains (Schmahmann, 2000). The cerebellum is thought to aid behaviour by learning from external input and forming internal models for automatic processing and more efficient behaviour (Balsters & Ramnani, 2011). Functional specialisation would therefore be dictated by the connectivity profile of a given cerebellar subregion (Guell et al., 2018). As a result, detrimental effects of grey matter loss should be region-specific and can affect a variety of different functional domains by virtue of poorer modulation of behaviour as the communication between cerebellum and neocortical and limbic regions is impaired (Jacobs et al., 2018).

Prior research and our findings suggest that age- and AD-related changes in multiple cerebellar subregions also impact multiple functional domains, especially those supporting cognitive processes (Gellersen et al., 2017). In posterior cerebellar regions, grey matter volume loss has been associated with performance on a variety of cognitive functions in healthy older adults and AD many of which co-localise with regions in our meta-analysis (Bernard & Seidler, 2014; Jacobs et al., 2018; Thomann et al., 2008). This can even be found when controlling for cerebral cortical or hippocampal volume, strongly suggesting an independent contribution to cognitive status in ageing and AD (Buhrmann et al., 2020; Lin, Chen, Tom, & Kuo, 2020).

A finding unique to the ageing as opposed to the AD data was a cluster in a cerebellar region of the DMN that may contribute to emotion processing (King et al., 2019). Despite its well established role in affective symptoms across a range of neuropsychiatric disorders (Schmahmann, 2004), we are only aware of one study that linked the volume of the same Crus II region and its cortical connectivity pattern with altered emotional processing in healthy older adults (Uwisengeyimana et al., 2020).

More common in the healthy ageing literature are positive associations between grey matter volume of the cerebellum and executive functioning, memory and language processing: long-term memory performance is related to larger total cerebellar volume and lobules VI and Crus II (Becker et al., 2015; Hafkemeijer et al., 2014; Koppelmans et al., 2017; Serra et al., 2017); posterior cerebellar volume, especially Crus II, is associated with scores on language tasks (Rodríguez-Aranda et al., 2016); and the most frequent association between cerebellar volume and cognition can be observed for tasks involving executive functions and processing speed, most notably in lobule VI and Crus I (Buhrmann et al., 2020; Koini et al., 2018; Liang & Carlson, 2019; Ruscheweyh et al., 2013; Uwisengeyimana et al., 2020; Zhang et al., 2013). These findings are in line with the functional mappings we used here and mostly dovetail with our results in that age-related grey matter loss was found in cognitive regions involved in DMN, FPN and attention networks. However, our findings only show marginal involvement of the most lateral Crus I/II regions which are associated with autobiographical memory recall. These regions may not have been prominent in our meta-analysis because they were not consistently affected across multiple studies. Alternatively, deficits in cognitive control may have been the driver of the association between cerebellar grey matter decline and long-term memory performance, as executive functioning has been found to be a major factor in explaining age-related decline in strategic retrieval (Gellersen, Trelle, Henson, & Simons, 2020; Trelle, Henson, Green, & Simons, 2017).

Although the healthy ageing studies included in our meta-analysis did not specifically relate cerebellar grey matter to cognition, older adults in these studies tended to exhibit deficits in cognitive domains subserved by those cerebellar regions that exhibited age-related grey matter loss. Prior findings are therefore in line with the functional mappings we used here and mostly dovetail with our results in that age-related grey matter loss was found in cognitive cerebellar regions involved in DMN, FPN and attention networks.

Prior work has also suggested that connectivity between the cerebellum and the cerebral cortex is reduced in ageing (Bernard & Seidler, 2014). This may explain recent data showing age differences in cerebellar engagement across cognitive domains including lobule VI and Crus I/II, the regions observed in our findings (Bernard et al., 2020). The authors hypothesise that fewer cerebellar resources reduce the support of automatic processes based on internal behavioural models (Bernard et al., 2020). If that is indeed the case and additional resources are required to compensate for the lack of automatic processing, this may be particularly demanding on executive functions, which may have to engage in compensatory processes. Moreover, underactivation of posterior regions situated in FPN (Bernard et al., 2020) and the grey matter loss observed here in the same areas may contribute to a reduction in efficiency of cerebellar recruitment during cognitive tasks. Without longitudinal data, it remains unclear whether these changes in cerebellar engagement and neocortical-cerebellar connectivity are a result of grey matter loss or whether the opposite is the case. Future studies might examine whether decreases in grey matter in the regions identified here are driving such changes in BOLD activation.

There is mixed evidence with respect to the vulnerability of the cerebellum to amyloid beta. Some argue that the cerebellum is mostly resistant to amyloid pathology until later stages of Alzheimer’s disease (Liang & Carlson, 2019), others counter that AD pathology affects the cerebellum even in early stages leading to both motor and cognitive deficits in mouse models (Hoxha et al., 2018; Jacobs et al., 2018). AD pathology is likely to be related to cerebellar atrophy, as amyloid beta correlates with grey matter in the cerebellum even among healthy older adults (Oh, Madison, Villeneuve, Markley, & Jagust, 2014) and cerebellar volume loss occurs in both sporadic and early-onset AD (Gellersen et al., 2017; Guo et al., 2016a; Jacobs et al., 2018). It has been suggested that one reason for prior absence of amyloid pathology in the cerebellum may be methodological limitations in staining techniques, rather than a true resistance (Jacobs et al., 2018). As a result, the cerebellum may have been under-researched and used as a reference region in PET imaging, precluding further findings of AD pathology in patients. Multiple reviews of the literature dovetail with our findings that the cerebellum still plays a role in AD (Gellersen et al., 2017; Jacobs et al., 2018; Lin et al., 2020).

Our functional mappings showed that AD-related grey matter loss occurred in right Crus I/II and lobule VI, which are on the cognitive extreme of the cognitive-motor gradient but show no preference along the gradient measuring task positivity, in line with their participation in both default mode and frontoparietal networks (Buckner et al., 2011; Guell et al., 2018). Previous studies on cerebellar grey matter volume and its association with cognition in AD have found deficits in those functions that map onto the regions we identified as most vulnerable to AD-related atrophy: language and attentional processes, as well as cognitive control of memory retrieval and other executive functions, including processing speed and working memory (Arnaiz & Almkvist, 2003; Baldaçara et al., 2011; Dos Santos et al., 2011; Hoche, Guell, Vangel, Sherman, & Schmahmann, 2018; Lin et al., 2020; Thomann et al., 2008). However, it is of note that there are also multiple occasions on which standard neuropsychological dementia screening tools of global cognition (such as the Mini Mental State Exam or the ADAS-Cog) are unrelated to cerebellar volume (Colloby, O’Brien, et al., 2014; Farrow et al., 2007; Lin et al., 2020; Möller et al., 2013). One explanation may be that cerebellar grey matter loss can still be compensated in earlier stages of the disease. Another reason that is likely is that these standard tools do not assess specific cognitive functions in depth, collapse scores across domains and therefore do not tap into the specific functions supported by the cerebellum.

Grey matter loss as observed in our meta-analysis may contribute to the altered connectivity between hippocampus, cerebral, and cerebellar DMN regions already seen in early AD, individuals with mild cognitive impairment (MCI) and even healthy older adults with subjective memory complaints (Bai et al., 2009; Greicius et al., 2004; Hafkemeijer et al., 2013; Serra et al., 2017). This may result in a reduction in efficient communication with neocortical and limbic regions. Indeed, patients with greater functional connectivity between the cerebellum and neocortex exhibited better cognitive performance (Liang & Carlson, 2019).

Even the strongest genetic risk factor for AD (the epsilon-4 allele of the apolipoprotein gene) has been shown to affect cerebellar communication with corticolimbic regions during memory tasks in young e4-carriers, which occurred in the absence of differences in cerebellar grey matter volume and did not translate to differences in associative memory performance between genetic groups (Matura et al., 2014). These functional effects in young e4-carriers bore a striking resemblance with the cluster in right Crus I/lobule VI that we identified here and may be a precursor to obvious grey matter and cognitive decline in later disease stages.

Longitudinal studies that could track the evolution of cerebellar volume and function across the AD-continuum are needed to better understand the role of the cerebellum in functional decline and its interplay with other brain areas. Differential subregional vulnerability to AD pathology or to normal ageing may be due to downstream influences resulting from cerebro-cerebellar synergism (Raz & Rodrigue, 2006; Sullivan & Pfefferbaum, 2006). Structural decline in the cerebellum may also have selective effects on distant neocortical regions (Limperopoulos, Chilingaryan, Guizard, Robertson, & Du Plessis, 2010). Indeed, neuroimaging studies have revealed covariation in grey matter volume throughout neocortical and cerebellar regions (Guo et al., 2016b; Hogan et al., 2011). Recent studies on longitudinal changes in cerebellar volume in AD have come to conflicting results, with one suggesting that cerebellar volume loss only contributes to disease progression in later stages from MCI to AD (Tabatabaei-Jafari, Walsh, Shaw, & Cherbuin, 2017), whereas another stipulates that the opposite is true (Lin et al., 2020). Both studies only consider total cerebellar volume and did not relate changes in grey matter to changes in cognition between baseline and follow-up. A mechanistic understanding of the cerebellum’s role in AD is therefore still lacking.

### 4.3 Limitations and future directions

It is surprising that we did not find clusters of significant age-related grey matter loss in anterior motor regions of the cerebellum. Prior evidence has shown volumetric decline of motor regions in normal ageing which correlates with motor deficits (Bernard et al., 2015; Bernard & Seidler, 2013; Hulst et al., 2015; Koppelmans et al., 2017; Liang & Carlson, 2019; Uwisengeyimana et al., 2020). However, some of these and other studies assessed in our literature search did not include coordinates of grey matter loss and could not be used in the present meta-analysis. Our analysis likely underestimates the extent of grey matter reductions in the cerebellum in healthy ageing and should not be interpreted as a demonstration of the absence of an age effect on cerebellar motor regions. Replication based on data from single subjects is an important next step.

It should be noted though, that this apparent limitation of the coordinate-based meta-analysis also presents an advantage. Our meta-analytic method does not require lobular boundaries as a constraint, which would be detrimental to the interpretation of functional implications given that anatomical borders do not translate neatly to boundaries between functional domains (King et al., 2019). These recent parcellations demonstrate that the mere knowledge of the lobule in which grey matter atrophy occurs may be too simplistic for the characterisation of function. Previous volumetry studies likely combine distinct functional territories which may obscure volume-function relationships. Especially those studies focusing on total grey matter volume cannot further our understanding of the precise role of regional cerebellar grey matter loss for specific cognitive changes in ageing or AD (Lin et al., 2020; Tabatabaei-Jafari et al., 2017).

As discussed above, prior studies and our findings converged to point to right regions of lobule VI and Crus I/II as particularly vulnerable to age, AD genetic risk and actual AD cases. AD may exacerbate the downward trajectory of this region already observed in healthy ageing shown here. Alternatively, the studies that found significant age-related grey matter loss in these regions may point out early declines that will later manifest as AD. Our data are limited in that they cannot test this possibility given the lack of longitudinal data in our analysis.

It is not clear to what extent cerebellar structural decline has clinical relevance for behavioural deficits in AD patients. As mentioned above, cerebellar deficits alone may not be sufficient to result in scores below the cut-off point for normal functioning in standard AD screenings because they are masked by the large number of test items that are insensitive to cerebellar function (Hoche et al., 2018). It is possible that correlations between cerebellar grey matter and affective-cognitive functioning in AD may be detected more readily with a more sensitive tool geared towards deficits caused by cerebellar decline, such as the novel Cerebellar Cognitive Affective/Schmahmann Syndrome Scale (Hoche et al., 2018). This may be an interesting avenue for future research.

Finally, our findings may be of relevance for studies aiming to determine optimal targets for neuromodulatory therapeutic approaches in ageing and AD. Non-invasive modulation of cerebellar regions has been shown to have network- and task-specific effects on connected neocortical regions (D’Mello, Turkeltaub, & Stoodley, 2017; Halko, Farzan, Eldaief, Schmahmann, & Pascual-Leone, 2014). Targeting cerebellar regions that are part of cortico-limbic networks most affected by AD through augmentative neuromodulation may bolster long-range connectivity.

### 4.4 Conclusions

Structural and functional analyses combined paint a picture of divergent and convergent patterns of cerebellar vulnerability to age and AD. The meta-analysis highlights vulnerability of posterior cerebellum to AD and ageing (Crus I/II, lobule VI), suggesting that some aspects of the age-related downward trajectory of grey matter may be exacerbated by AD pathology. The strict right lateralisation of AD atrophy compared to bilateral effects in healthy ageing was striking. Notably, besides being located in the same lobules, the subregions of structural atrophy for AD and ageing were mainly non-overlapping. Nonetheless, regions with vulnerability to AD and ageing exhibited similar functional profiles: they participated in the same large-scale discrete functional networks, were involved in working memory, attention and language processing, and were located in similar positions along major functional gradients (cognitive vs. motor and degree of task focus). Despite these similarities, differences along Gradient 3 allowed for the observation of age-AD divergence along alternative dimensions of cerebellar functional hierarchies as exemplified by different preferences along the tertiary cerebellar gradient (lateralisation in cognitive territories) and showed that only regions with age-related grey matter decline were involved in affective processes. These findings show subtler differences in the functional fingerprint of regions impacted by AD and ageing that the standard categorical parcellations of brain networks was not able to detect. Our study provides an unprecedented characterisation of structural and functional localisation differences and similarities in cerebellar atrophy between ageing and AD. They also show how the use of multiple functional maps of the cerebellum is synergistic in interpreting the implications of regional grey matter loss. More research is needed to better understand 1) how age and pathology impact cerebellar computational processes, 2) how cerebellar interactions with other brain regions may influence cognitive decline or preservation in healthy ageing and AD, 3) and the role of subregional cerebellar grey matter loss in the neuropsychology of AD.

## Data Availability

Data were taken from the primary studies cited in this manuscript and are therefore freely available in the original papers.

