## Appendix A for "Differential vulnerability of the cerebellum in healthy ageing and Alzheimer’s disease"

**Appendix A – Details of systematic literature search**

**Table A1. Search terms for the systematic literature search.**

| **Date Searched** | **Database** | **Search Terms and Restrictions** | ***N* identified studies** |
| --- | --- | --- | --- |
| October 1st 2019 | Pubmed | (*ag*ing* OR *elderly* OR *older adults* OR *age-related*) AND (*voxel-based morphometry* OR *VBM* OR *grey matter* OR *gray matter*) in **Title/Abstract**; filter: human | 1766 |
|  | PsycInfo | (*ag*ing* OR *elderly* OR *older adults* OR *age-related*) AND (*voxel-based morphometry* OR *VBM* OR *grey matter* OR *gray matter*) in **Abstract** (filter: human, peer-reviewed journals, English) | 1078 |
| Unique records | | | 2031 |

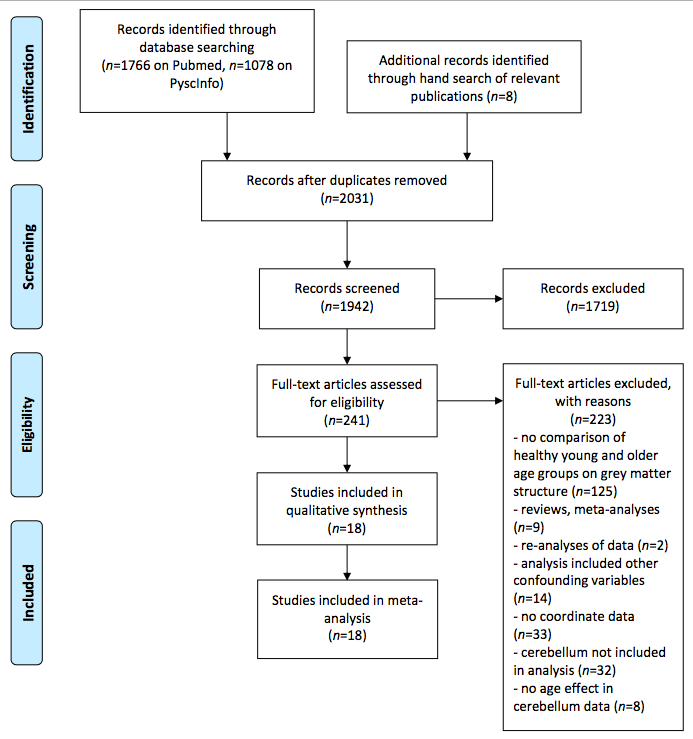

**Figure A1. PRISMA flowchart of the study selection procedure.**
