## Appendix B for "Differential vulnerability of the cerebellum in healthy ageing and Alzheimer’s disease"

**Appendix B – Study Characteristics**

**Table B1. Summary of studies included in the meta-analysis of cerebellar grey matter loss in healthy ageing.**

| *Studies using direct comparisons of age groups* | | | | | | | | |  | |
| --- | --- | --- | --- | --- | --- | --- | --- | --- | --- | --- |
| **Author, year** | ***N***  **(% female)** | **Age ± *SD* (range)** | **Software** | **Preprocessing** | **Nuisance covariates; significance level** | **Foci for age effect**  **(MNI)** | **Cognitive deficits in older vs. younger adults** | | **Additional Notes** | |
| Maguire & Frith, 2003  [1] | YA  12  (50)  OA  12 (50) | YA  32±4  (23-39)  OA  75±5  (67-80) | SPM99 | Optimised  Unmodulated | *p*<.05 corrected | -47 -77 -44 | ↓ Autobiographical fact retrieval  = Memory for autobiographical events  = Memory for public events  = General knowledge | |  | |
| Steffener, Brickman, Rakitin, Gazes, & Stern, 2009  [2] | YA  37  (22)  OA  15  (53) | YA  25±4  OA  75±7 | SPM5 | Optimised  Modulated | Normalised whole brain volume  *p<*.005 uncorrected | 6 -74 -12 | ↑ Reaction times for larger set sizes in a Sternberg working memory task | |  | |
| Antonova et al., 2009  [3] | YA  10  OA  10 | YA  24±2  (20-26)  OA  72±5  (64-79) | SPM2 | Optimised  Modulated | NA  *p*<.05 FWE | -22 -58 -32  3 -75 -17  39 -69 -25  27 -57 -29  26 -48 -30^a^ | ↓ Accuracy for spatial location of objects in a spatial scene | |  | |
| Bauer, Gebhardt, Gruppe, Gallhofer, & Sammer, 2012  [4] | YA  18  OA  18 | YA  24±2  (19-28)  OA  60±6  (54-77) | SPM8 | Unified  DARTEL  Modulated | TIV  *p*<.05 FWE | 30 -66 -30  -23 -75 -23 | ↑ Reaction times during a location priming task  = Errors in location priming task | |  | |
| Kalpouzos, Persson, & Nyberg, 2012  [5] | YA  16  (50)  OA  20  (100) | YA  25  (21-39)  OA  61  (52-69) | SPM5 | Unified  Modulated | *p*<.001 FEW | -26 -35 -38 | Unclear | |  | |
| **Author, year** | ***N***  **(% female)** | **Age ± *SD* (range)** | **Software** | **Preprocessing** | **Nuisance covariates; significance level** | **Foci for age effect**  **(MNI)** | **Cognitive deficits in older vs. younger adults** | **Additional Notes** | | |
| Bauer, Sammer, & Toepper, 2018)  [6] | YA  35  (57)  OA  35  (49) | YA  27±5  (20-35)^b^  OA  61±7  (50-80)^b^ | SPM12 | Pipeline through CAT12  Modulated | TIV, gender, years of education  *p*<.05 FWE | 29 -77 -39  -32 -72 -41  -24 -45 -41  -45 -53 -45 | ↑ Errors in a high load working memory task (Corsi Block-Tapping) |  | | |
| *Studies using age as continuous variable* | | | | | | | |  | | |
| Good et al., 2001  [7] | 465  (43) | 30^c^  (17-79) | SPM99 | Optimised  Modulated | TIV, linear and nonlinear age effects  *p*<.05 corrected for multiple comparisons | 31 -90 -34 | NA |  | | |
| Alexander et al., 2006  [8] | 26  (42) | 51±16  (22-77) | SPM2 | Optimised  Modulated | TIV  *z* ≥ -2 | 38 -89 -26^a^ | No cognitive decline (MMSE>27 in all participants) | Scaled subprofile model;  only regions with negative associations with age | | |
| Abe et al., 2008  [9] | 73  (100) | 39.2±14.9  (22-70) | SPM2 | Optimised  Modulated | TIV  *p*<.05 FWE | 26 -86 -32  -24 -88 -32 | NA |  | | |
| Kalpouzos et al., 2009  [10] | 45  (53) | 49±18  (20-83) | SPM2 | Optimised  Modulated | *p*<.01 FDR  minimum cluster size *k*=20 | -46 -46 -41  36 -43 -44 | NA |  | | |
| Bergfield et al., 2010  [11] | 29^d^  (62) | 48±19  (23-84) | SPM2 | Optimised  Modulated | TIV  *z* ≥ -2 | 52 -47 -29^a^ | NA | Scaled subprofile model; only regions with negative associations with age | | |
| **Author, year** | ***N***  **(% female)** | **Age ± *SD* (range)** | **Software** | **Preprocessing** | **Nuisance covariates; significance level** | **Foci for age effect**  **(MNI)** | **Cognitive deficits in older vs. younger adults** | **Additional Notes** | | |
| Draganski et al., 2011  [12] | 26  (27) | 52  (18-85) | SPM8 | Unified,  DARTEL  Modulated | Gender, TIV  *p*<.05 FWE | -36 -58 -31  39 -58 -32 | NA |  | | |
| Salami, Eriksson, & Nyberg, 2012  [13] | 292  (52) | 60±13  (25-80) | SPM8 | Unified  DARTEL | Age and  square of orthogonalised age  TIV  *p*<.05 FDR | 35 -60 -30 | ↓ Names recalled during face-name association task | Data from Betula Prospective Study | | |
| Ziegler et al., 2012  [14] | 547  (56) | 48±17 | SPM8 | Optimised  Modulated | Scanning site, linear and quadratic age effects  *p*<.05 FWE | 15 -54 -17  20 -61 -57  -20 -55 -18  -20 -79 -35  6 -72 -44 | NA | Non-linear negative age effect | | |
| Thürling et al., 2014)  [15] | 34  (56) | 42±16  (21-74) | SPM8 | SUIT normalised  Modulated | TIV  *p<*.05 FWE | -8 -67 -23  18 -74 -29  -13 -48 -43  -30 -61 -20  -35 -70 -41  -4 -75 -20  35 -73 -39  -30 -85 -25  49 -64 -27  -30 -52 -18  32 -65 -18  -13 -66 -42  -41 -76 -39  -35 -61 -50  8 -67 -25  -41 -73 -29  36 -65 -25  -11 -73 -39  49 -55 -29 | ↓ Storage and extinction of visual threat eyeblink responses; correlated with total cerebellar volume | ROI analysis of the cerebellum; coordinates in SUIT space | | |
| **Author, year** | ***N***  **(% female)** | **Age ± *SD* (range)** | **Software** | **Preprocessing** | **Nuisance covariates; significance level** | **Foci for age effect**  **(MNI)** | **Cognitive deficits in older vs. younger adults** | **Additional Notes** | | |
| Dickie et al., 2015  [16] | 80  (50) | 43^e^  (25-64) | FSL-VBM | Optimised  Modulated | NA  *p*<.05 FDR | 20 -66 -28  -48 -60 -30 | NA | Included only results from permutation testing with 20,000 iterations | | |
| Yu, Korgaonkar, & Grieve, 2017  [17] | 438^f^  (58) | 32±19  (7-86) | SPM8 | SUIT normalised  Modulated | TIV, gender,  scanning site  *p*<.05 FWE | -37 -39-39  51 -55 -31  -3 -80 -28  -31 -82 -38  34 -83 -38  -7 -65 -44  17 -31 -21  2 -67 -14  10 -66 -41 | NA | | | ROI analysis of the cerebellum; only data pertaining to GM loss, not preservation, is included |
| Hu et al., 2018  [18] | 149  (56) | 32±12  (18-72) | SPM8 | DARTEL  Modulated | NA  *p<*.05 FWE | -35 -82 -39  -8 -70 -14  -39 -40 -42  38 -40 -42  38 -79 -39 | ↑ Reaction time in a stop signal task | | |  |

^a^Coordinates transformed from Talairach & Tournaux to MNI

^b^Numbers based on mean of high and low performing cognitive group

^c^Mean of four medians from four groups of participants (right-handed females, right-handed males, left-handed females, left-handed males)

^d^Only data from Group 1 included because Group 2 was assessed in Alexander et al. (2006)

^e^Median age

^f^Total study sample size minus 41 subjects who were excluded from the VBM analysis

Abbreviations. CAT: computational anatomy toolbox. DARTEL: diffeomorphic anatomical registration trough exponentiated lie algebra; FDR: false discovery rate. FWE: familywise error; FSL: functional magnetic imaging of the brain (FMRIB) software library; GM: grey matter. MA: middle-aged adults. MNI: Montreal Neurological Institute; N: sample size; NA: not applicable; OA: older adults. ROI: region of interest. SD: Standard Deviation. SPM: Statistical Parametric Mapping. SUIT: spatially unbiased infratentorial template. TIV: total intracranial volume. VBM: voxel-based morphometry. YA: younger adults.

**Table B2. Study characteristics of records included in the coordinate-based meta-analysis.** Studies are listed according to the degree of cognitive impairment going from least to most severe as measured using the MMSE. Note that this table is based on Gellersen et al. (2017)*: <https://jnnp.bmj.com/content/88/9/780.full#DC1>.

| **Authors** | **Notes on diagnosis** | ***N* patients (% female)** | ***N* controls (% female)** | **Age patients**  **± *SD*** | **Age controls**  **± *SD*** | ***p*-value age difference** | **MMSE or ACE patients**  **± *SD*** | **Disease Duration**  **(years ± *SD*)** | **Coordinates (MNI)** | **Relationship between cerebellar grey matter and cognition and clinical ratings**  **(NA if no such analysis was carried out)** | **Cognitive deficits in patients vs. controls** |
| --- | --- | --- | --- | --- | --- | --- | --- | --- | --- | --- | --- |
| Farrow et al., 2007  [19] | Probable AD based on NINCDS-ADRDA criteria | 7 | 11 | 78±7 | 71±4 | .014 | 25±4 (MMSE) | 4 | 25 -40 -29  -24 -36 -29 | No correlation between ADAS-TES/MMSE and cerebellar GM | ↓ MMSE, ADAS-TES |
| Mazère et al., 2008  [20] | Probable based on NINCDS-ADRDA criteria | 8 (63) | 8 (75) | 80±7 | 74±3 | NS | 24±2 (MMSE) | NA | -44 -65 -42  27 -66 -11 | NA | ↓ MMSE |
| Ossenkoppele et al., 2015  [21] | Subgroup of AD patients defined as typical AD based on NIA-AA criteria; biomarker confirmed | 58 (39) | 61 (38) | 64±9 | 64±8 | NS | 23±4 (MMSE) | NA | -39 -82 -33  46 -73 -36 | NA | ↓ MMSE  47% of patients with memory impairment  7% with executive functioning |
| Canu et al., 2011  [22] | Based on NINCDS-ADRDA criteria | 17 (82) | 13 (46) | 77±6 | 73±7 | NS | 21±5 (MMSE) | NA | 42 -59 -25  32 -64 -36  -29 -70 -39  -36 -67 -32 | NA | ↓ MMSE |
| Möller et al., 2013  [23] | Subgroup of late-onset probable AD based on NINCDS-ADRDA criteria | 120 (46) | 71 (50) | 72±5 | 71±4 | NS | 21±5 (MMSE) | NA | 33 -60 -27  30 -69 -38  12 -61 -23  26 -49 -47  -26 -48 -45  -34 -48 -45  -30 -42 -42  10 -67 -36 | No correlation between MMSE and cerebellar GM | ↓ MMSE  ↓ RAVLT immediate and delayed  ↓ Trail Making Test A and B |
| Colloby, O’Brien, & Taylor, 2014  [24] | Probable AD based on NINCDS-ADRDA criteria | 47 | 39 | 79±9 | 77±6 | NS | 21±4 (MMSE) | NA | -33 -43 -24  42 -43 -26 | No correlation between MMSE and cerebellar GM | ↓ MMSE  ↓ CAMCOG |
| **Authors** | **Notes on diagnosis** | ***N* patients (% female)** | ***N* controls (% female)** | **Age patients**  **± *SD*** | **Age controls**  **± *SD*** | ***p*-value age difference** | **MMSE or ACE patients**  **± *SD*** | **Disease Duration**  **(years ± *SD*)** | **Coordinates (MNI)** | **Relationship between cerebellar grey matter and cognition and clinical ratings**  **(NA if no such analysis was carried out)** | **Cognitive deficits in patients vs. controls** |
| Canu et al., 2012  [25] | Subgroup of late-onset probable AD based on NINCDS-ADRDA criteria | 24 (67) | 24 (71) | 78±5 | 76±4 | NS | 21±4 (MMSE) | 4±2 | 33 -75 -28 | NA | ↓ MMSE  ↓ RCFT delayed  ↓ RAVLT immediate and delayed  ↓ Trail Making Test |
| Toniolo et al., 2018 [26] | Probable AD based on NINCDS-ADRDA criteria | 53 (66) | 34 (50) | 75±6 | 69±7 | NS | 20±3 (MMSE) | NA | 19 -35 -19  39 -63 -23  -4 -51 -26 | Correlation between anterior and posterior cerebellar grey matter volume and Copy of drawings test; no correlation with MMSE, short-term memory, long-term memory, word fluency, language or executive functions. | ↓ MMSE  ↓ RAVLT immediate and delayed  ↓ Phonological verbal fluency  ↓ Digit span forward and backward  ↓ Copy of drawings and drawings with landmarks  ↓ Raven’s progressive matrices  ↓ Corsi blocking task |
| Lehmann et al., 2011  [27] | Probable AD based on NINCDS-ADRDA criteria; typical AD presentation. | 30 (53) | 50 (66) | 69±9 | 63±10 | <.005 | 19±5 (MMSE) | 5 | 8 -49 -30 | NA | ↓ MMSE |
| Serra et al., 2014  [28] | Probable AD based on NINCDS-ADRDA criteria | 48 (35) | 20 (65) | 71±6 | 70±6 | NS | 19±3 (MMSE) | 4±3 | -12 -86 -24 | NA | ↓ MMSE  ↓ RAVLT immediate and delayed  ↓ RCFT delayed  ↓ Short story test  ↓ Corsi blocking task  ↓ Phonological word fluency  ↓ Card sorting test  ↓ Raven’s progressive matrices |
| **Authors** | **Notes on diagnosis** | ***N* patients (% female)** | ***N* controls (% female)** | **Age patients**  **± *SD*** | **Age controls**  **± *SD*** | ***p*-value age difference** | **MMSE or ACE patients**  **± *SD*** | **Disease Duration**  **(years ± *SD*)** | **Coordinates (MNI)** | **Relationship between cerebellar grey matter and cognition and clinical ratings**  **(NA if no such analysis was carried out)** | **Cognitive deficits in patients vs. controls** |
| Guo et al., 2016  [29] | Probable AD based on NINCDS-ADRDA criteria | 34 (44) | 34 (53) | 62±6 | 64±5 | NS | NA (MMSE) | 3±3 | -32 -72 -29  -31 -60 -19  27 -71 -28  27 -76 -26 | NA | ↓ RAVLT  ↓ RCFT  ↓ Doors and people test |
| Raji et al., 2009  [30] | Probable AD based on NINCDS-ADRDA criteria | 33 (33) | 169 (57) | 83±5 | 78±4 | .001 | 76±13^a^ (MMSE) | NA | -24 -33 -31  28 -33 -34  1 -37 -20 | NA | ↓ MMSE |
| Ahmed et al., 2019 [31] | Probable AD based on NINCDS-ADRDA criteria | 16 (38) | 19 (32) | 60±6 | 63±7 | NS | 62±16 (ACE-III) | 4±2 | -52 -58 -46  42 -52 -58 | NA | ↓ ACE-III |

^a^Use of modified MMSE.

Abbreviations. ACE-III: Addenbrookes Cognitive Examination – Version 3. AD: Alzheimer’s disease. ADAS-TES: Alzheimer's Disease Assessment Scale Total Error Score. CAMCOG: Cambridge Cognitive Examination. MMSE: Mini Mental State Exam. MNI: Montreal Neurological Institute. NA: not available. NIA-AA: National Institute on Aging-Alzheimer’s Association. NINCDS-ADRDA: National Institute of Neurological and Communicative Disorders and Stroke and Alzheimer's Disease and Related Disorders Association. NS: not significant. RAVLT: Rey Auditory Verbal Learning Test. RCFT: Rey Complex Figure Test. SD: Standard Deviation.
