## Appendix C for "Differential vulnerability of the cerebellum in healthy ageing and Alzheimer’s disease"

**Appendix C – Testing Gradient Differences in Ageing and Alzheimer’s Disease**

**a.**

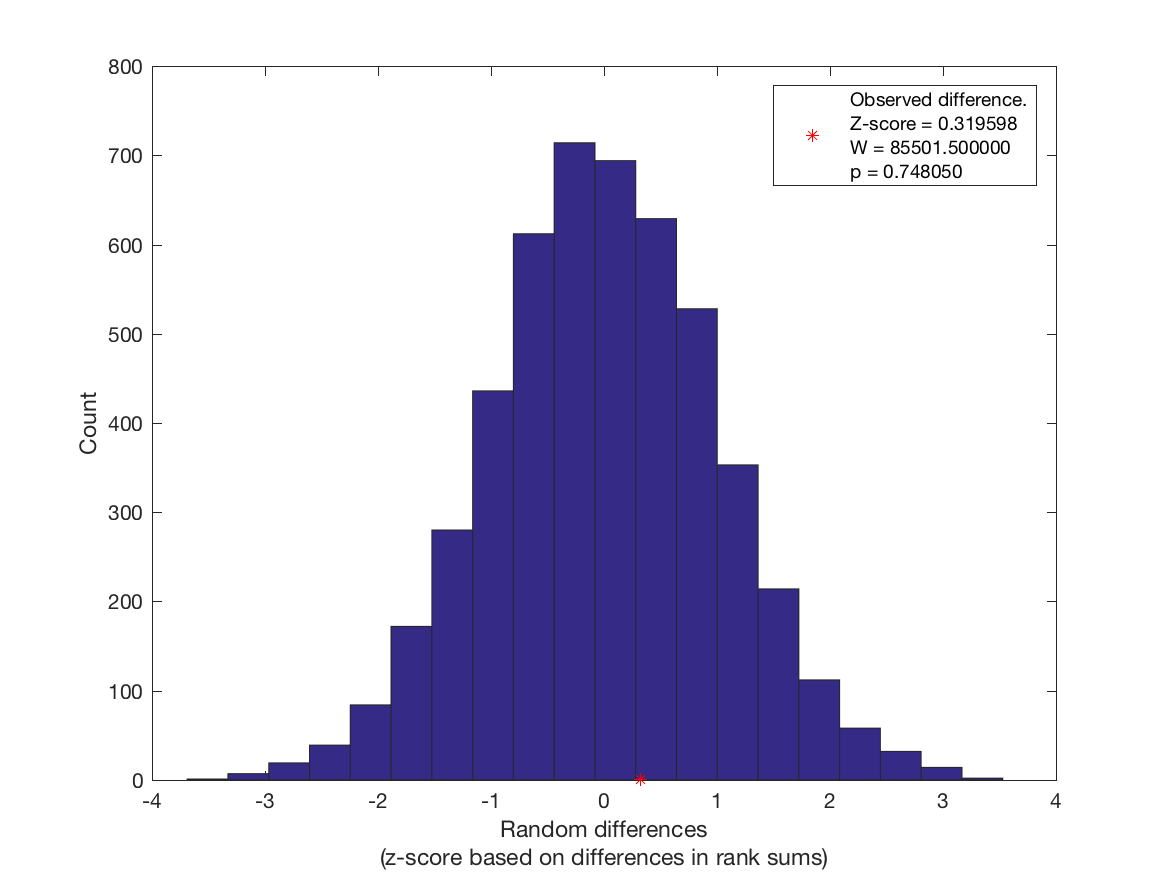
 **b.**

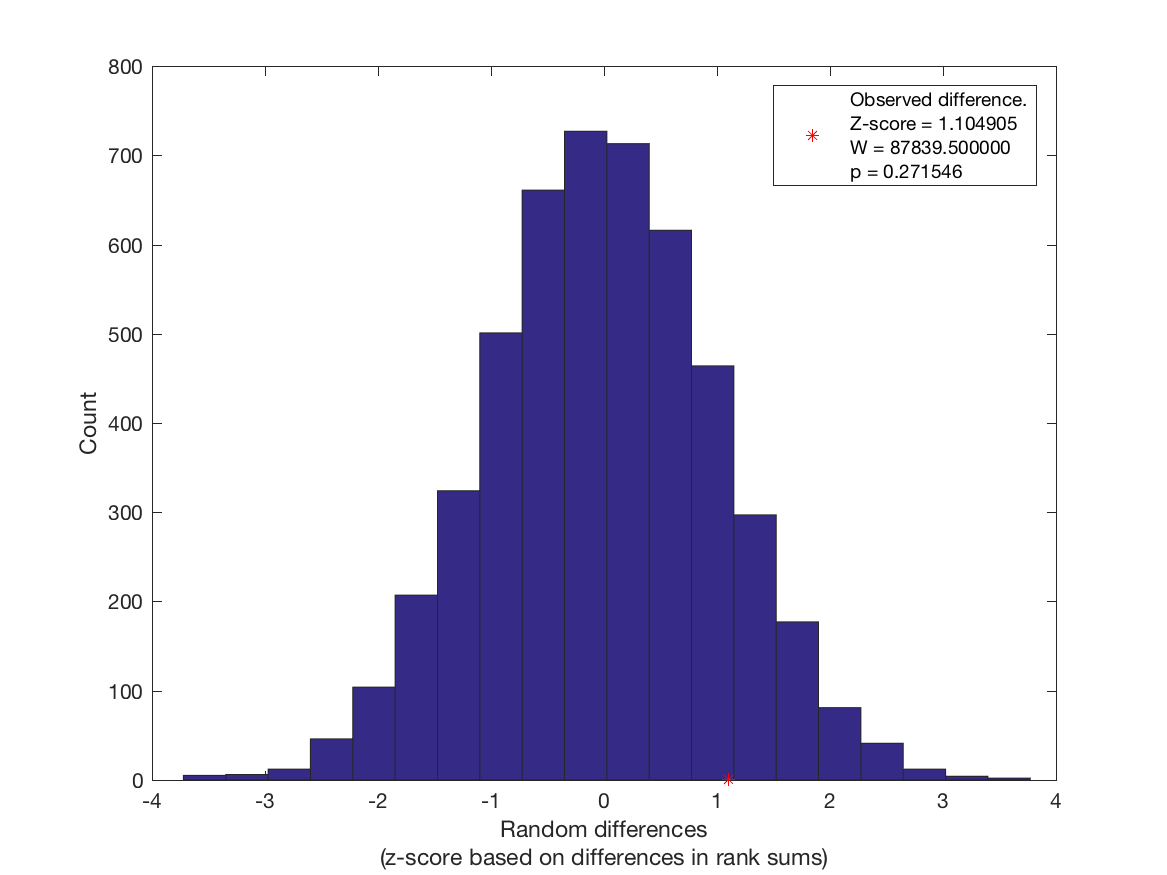

**c.**

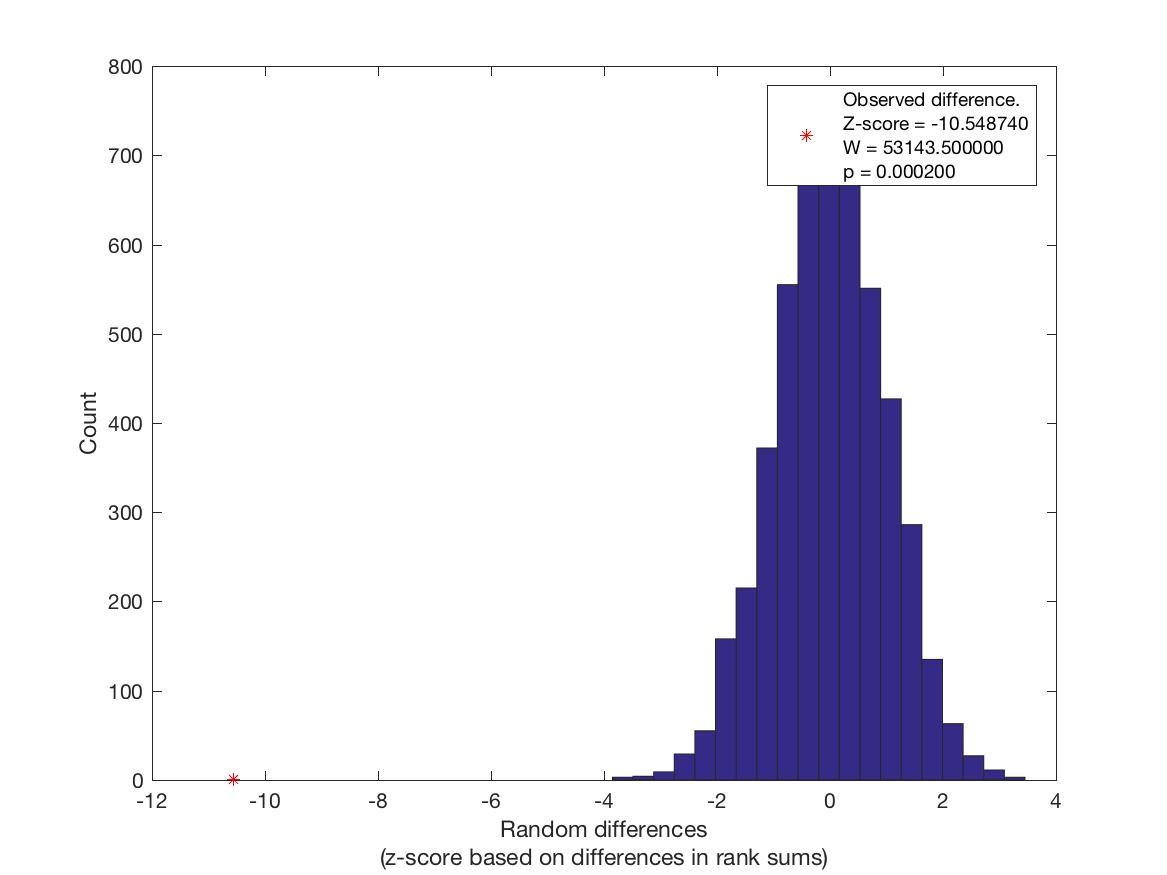

**Figure C1. Results of the permutation analysis to assess differences in values of the functional gradients for AD- and age-related grey matter decline. Observed differences between healthy ageing and AD gradient values were computed using a Wilcoxon rank sum test and a subsequent conversion of ranks to *z*-scores. Positive differences indicate higher values for the healthy ageing compared to the AD group. Observed differences are marked with a red asterisk. Random differences were calculated based on random allocation of a given gradient value to the ageing or AD group. a. Gradient 1 values. B. Gradient 2 values. c. Gradient 3 values.**
