## Appendix D for "Differential vulnerability of the cerebellum in healthy ageing and Alzheimer’s disease"

**Appendix D – Results of Robustness Tests**

Results of each jackknife analysis can be found in Table D1. A count of the occurrences of cluster survival, size changes, and peak location shifts revealed that, for the case of healthy ageing, there was no case in which a one study removed analysis changed the label of the peak voxel of a cluster (Table D2). Only slight shifts in peak voxel locations were observed. The jackknife procedure showed that clusters 1 and 2 in right Crus I/II were robust to the removal of any one study. Only unsubstantial changes in size and extent of these clusters occurred. Cluster 3 in posterior Crus II did not survive the removal of the study by Hu et al. [1] (*n*=149 subjects, *k*=5 foci), while cluster 4 in vermal/right lobule VI did no longer emerge when the study by Antonova et al. [2] (*n*=20, *k*=5 foci) was removed. Both cluster 3 and 4 remained stable in all other instances, indicating robustness in 94% of analyses. Finally, cluster 5 in anterior Crus I/II remained in 83% of all analyses but did not survive correction for multiple comparisons upon removal of studies that were the main contributors to this cluster [1–4]. These also tended to be studies with a larger number of foci.

The two clusters for the AD meta-analysis remained present 85% of all jackknife analyses (Table D2), with two studies each resulting in either cluster 1 or cluster 2 not surviving the FEW *p*<.05 threshold (but it was never the case that both clusters disappeared).

In five instances, the two clusters merged into one larger cluster. Changes in the coordinate of the peak location of the cluster did occur but this change never affected the label of this location. In five cases, the cluster size was reduced (removal of studies by [5–9]).

Figures D1-3 show cerebellar flatmaps and gradient maps for all one-study-removed analyses to visualise the changes in structural age and AD effects and their localisation onto functional gradients caused by the removal of each individual study. We further assessed the effect of the jackknifing procedure on mean gradient values and tested whether the difference in gradient values we observed between our full ageing analysis with all studies included and our previous AD meta-analysis was robust to jackknifing (Figure D4 for a distribution of mean gradient values). Running the permutations to test for a difference in gradient values for each of the 13 AD and 18 ageing jackknife analyses revealed that on average, there was no difference in Gradient 1 values between healthy ageing and AD cases (*Mean z-*score based on Wilcoxon rank sum=.145, 95% *CI* [-.682, .973]; *Cohen’s U_1_*=.215, 95% *CI* [.182, .248]). Likewise, there was no difference in Gradient 2 values (*Mean z*=.928, 95% *CI* [.148, 1.707]; *Cohen’s U_1_*=.104, 95% *CI* [.071, .137]). The distributions of Gradient 3 values in AD and healthy ageing were significantly non-overlapping across one-study removed analyses (*Mean z*=-9.681, 95% *CI* [-10.381, -8.981]; *Cohen’s U_1_*=.337, 95% *CI* [.308, .367]).

**Table D1. Results of the jackknifing procedure. One study at a time was excluded to test robustness of the clusters from the main analysis.**

| **Study Removed** | **Number of foci** | **Number of subjects** | **Min cluster size (mm^3^) by ALE algorithm** | **Cluster number (by size; ordered according to correspondence to main analysis cluster)^a^** | **Change in cluster size?** | **Change in peak**  **coordinate by >2 in any direction?** | **Change in label of peak coordinates?** | **Extent**  **(mm^3^)** | **Cluster coordinates** |
| --- | --- | --- | --- | --- | --- | --- | --- | --- | --- |
| *Healthy ageing* | | | | | | | | | |
| None | 64 | 2441 | 520 | 1 | NA | NA | NA | 1504 | (24 -90 -42) to (40 -72 -30) centered at (33 -81 -37) |
|  |  |  |  | 2 |  |  |  | 1464 | (28 -70 -34) to (42 -56 -22) centered at (35 -63 -29) |
|  |  |  |  | 3 |  |  |  | 1288 | (-46 -86 -44) to (-28 -68 -34) centered at (-35 -77 -40) |
|  |  |  |  | 4 |  |  |  | 1112 | (-10 -78 -22) to (8 -66 -10) centered at (1 -72 -15) |
|  |  |  |  | 5 |  |  |  | 808 | (-48 -54 -46) to (-34 -36 -36) centered at (-40 -42 -41) |
| Abe et al., 2008  [11] | 62 | 2368 | 488 | 3 | ↓ | No | No | 1160 | (28 -90 -42) to (40 -72 -32) centered at (34 -80 -38) |
|  |  |  |  | 1 | ↑ | No | No | 1472 | (28 -70 -34) to (42 -56 -22) centered at (35 -63 -29) |
|  |  |  |  | 2 | = | No | No | 1288 | (-46 -86 -44) to (-28 -68 -34) centered at (-35 -77 -40) |
|  |  |  |  | 4 | = | No | No | 1112 | (-10 -78 -22) to (8 -66 -10) centered at (1 -72 -15) |
|  |  |  |  | 5 | = | No | No | 808 | (-48 -54 -46) to (-34 -36 -36) centered at (-40 -42 -41) |
| Alexander et al., 2006  [12] | 63 | 2415 | 520 | 1 | ↓ | No | No | 1464 | 24 -90 -42) to (40 -72 -30) centered at (33 -81 -37) |
|  |  |  |  | 2 | = | No | No | 1464 | (28 -70 -34) to (42 -56 -22) centered at (35 -63 -29) |
|  |  |  |  | 3 | = | No | No | 1288 | (-46 -86 -44) to (-28 -68 -34) centered at (-35 -77 -40) |
|  |  |  |  | 4 | = | No | No | 1112 | (-10 -78 -22) to (8 -66 -10) centered at (1 -72 -15) |
|  |  |  |  | 5 | = | No | No | 808 | (-48 -54 -46) to (-34 -36 -36) centered at (-40 -42 -41) |
| Antonova et al., 2009  [2] | 59 | 2421 | 504 | 1 | = | No | No | 1504 | (24 -90 -42) to (40 -72 -30) centered at (33 -81 -37) |
|  |  |  |  | 3 | ↓ | No | No | 920 | (28 -68 -34) to (42 -56 -24) centered at (35 -62 -29) |
|  |  |  |  | 2 | ↑ | Yes | No | 1312 | (-48 -86 -44) to (-28 -68 -34) centered at (-35 -77 -49) |
| Cluster 5 from main analysis does not survive | | | | 5 | NA | NA | NA | NA | NA |
|  | | | | 4 | = | No | No | 808 | (-48 -54 -46) to (-34 -36 -36) centered at (-40 -42 -41) |
| Bauer et al., 2012  [13] | 62 | 2405 | 504 | 1 | = | No | No | 1504 | (24 -90 -42) to (40 -72 -30) centered at (33 -81 -37) |
|  |  |  |  | 4 | ↓ | No | No | 1040 | (28 -70 -34) to (42 -56 -22) centered at (36 -62 -28) |
|  |  |  |  | 2 | ↓ | No | No | 1312 | (-46 -86 -44) to (-28 -68 -34) centered at (-35 -77 -40) |
|  |  |  |  | 3 | = | No | No | 1112 | (-10 -78 -22) to (8 -66 -10) centered at (1 -72 -15) |
|  |  |  |  | 5 | = | No | No | 808 | (-48 -54 -46) to (-34 -36 -36) centered at (-40 -42 -41) |
| Bauer et al., 2018  [14] | 60 | 2371 | 464 | 3 | ↓ | No | No | 1152 | (24 -90 -42) to (40 -72 -30) centered at (33 -82 -37) |
|  |  |  |  | 1 | ↑ | No | No | 1520 | (26 -70 -34) to (42 -56 -22) centered at (35 -63 -29) |
|  |  |  |  | 4 | ↓ | Yes | No | 736 | (-48 -86 -44) to (-28 -74 -34) centered at (-35 -81 -39) |
|  |  |  |  | 2 | ↑ | No | No | 1176 | (-10 -78 -22) to (8 -66 -10) centered at (1 -72 -15) |
| Note that cluster 5 from the original analysis split into two sub-clusters in this analysis | | | | 5 | ↓ | Yes | No | 616 | (-46 -46 -44) to (-34 -36 -36) centered at (-39 -40 -41) |
|  | | | | 6 | ↓ | Yes | No | 472 | (46 -58 -34) to (54 -52 -26) centered at (50 -55 -30) |
| Bergfield et al., 2009  [15] | 63 | 2412 | 528 | 1 | = | No | No | 1504 | (24 -90 -42) to (40 -72 -30) centered at (33 -81 -37) |
|  |  |  |  | 2 | ↑ | No | No | 1472 | (28 -70 -34) to (42 -56 -22) centered at (35 -63 -29) |
|  |  |  |  | 3 | = | No | No | 1288 | (-46 -86 -44) to (-28 -68 -34) centered at (-35 -77 -40) |
|  |  |  |  | 4 | = | No | No | 1112 | (-10 -78 -22) to (8 -66 -10) centered at (1 -72 -15) |
|  |  |  |  | 5 | = | No | No | 808 | (-48 -54 -46) to (-34 -36 -36) centered at (-40 -42 -41) |
| Dickie et al., 2015  [16] | 62 | 2361 | 512 | 1 | ↑ | No | No | 1512 | (24 -90 -42) to (40 -72 -30) centered at (33 -81 -37) |
|  |  |  |  | 2 | ↑ | No | No | 1496 | (28 -70 -34) to (42 -56 -22) centered at (35 -63 -29) |
|  |  |  |  | 3 | ↑ | No | No | 1336 | (-48 -86 -44) to (-28 -68 -34) centered at (-35 -77 -40) |
|  |  |  |  | 4 | ↑ | No | No | 1176 | (-10 -78 -22) to (8 -66 -10) centered at (1 -72 -15) |
|  |  |  |  | 5 | = | No | No | 808 | (-48 -54 -46) to (-34 -36 -36) centered at (-40 -42 -41) |
| Draganski et al., 2011  [17] | 62 | 2415 | 496 | 1 | = | No | No | 1504 | (24 -90 -42) to (40 -72 -30) centered at (33 -81 -37) |
|  |  |  |  | 4 | ↓ | No | No | 1088 | (28 -70 -32) to (40 -58 -22) centered at (34 -64 -28) |
|  |  |  |  | 2 | ↑ | No | No | 1312 | (-48 -86 -44) to (-28 -68 -34) centered at (-35 -77 -40) |
|  |  |  |  | 3 | = | No | No | 1112 | (-10 -78 -22) to (8 -66 -10) centered at (1 -72 -15) |
|  |  |  |  | 5 | = | No | No | 808 | (-48 -54 -46) to (-34 -36 -36) centered at (-40 -42 -41) |
| Good et al., 2001  [18] | 63 | 1976 | 512 | 4 | ↓ | Yes | No | 992 | (28 -86 -42) to (40 -72 -36) centered at (34 -78 -39) |
|  |  |  |  | 1 | ↓ | No | No | 1472 | (28 -70 -34) to (35 -63 -29) centered at (36 -60 -30) |
|  |  |  |  | 2 | ↑ | No | No | 1312 | (-48 -86 -44) to (-28 -68 -34) centered at (-35 -77 -40) |
|  |  |  |  | 3 | ↓ | No | No | 1112 | (-10 -78,-22) to (8,-66,-10) centered at (1 -72 -15) |
|  |  |  |  | 5 | = | No | No | 808 | (-48 -54 -46) to (-34 -36 -36) centered at (-40 -42 -41) |
| Hu et al., 2018  [1] | 59 | 2293 | 536 | 2 | ↓ | No | No | 1128 | (24 -90 -42) to (40 -72 -30) centered at (33 -81 -37) |
|  |  |  |  | 1 | ↑ | No | No | 1544 | (26 -70 -34) to (42 -56 -22) centered at (35 -63 -29) |
| Cluster 3 from the original analysis does not survive | | | | |  |  |  |  |  |
|  |  |  |  | 3 | ↓ | No | No | 840 | (-4 -78 -22) to (8 -66 -10) centered at (3 -73 -15) |
| Cluster 5 from the original analysis does not survive | | | | |  |  |  |  |  |
| Kalpouzos et al., 2009  [19] | 62 | 2396 | 520 | 1 | = | No | No | 1504 | (24 -90 -42) to (40 -72 -30) centered at (33 -81 -37) |
|  |  |  |  | 2 | ↑ | No | No | 1472 | (28 -70 -34) to (42 -56 -22) centered at (35 -63 -29) |
|  |  |  |  | 3 | ↑ | No | No | 1312 | (-46 -86 -44) to (-28 -68 -34) centered at (-35 -77 -40) |
|  |  |  |  | 4 | = | No | No | 1112 | (-10 -78 -22) to (8 -66 -10) centered at (1 -72 -15) |
| Cluster 5 from the original analysis does not survive | | | | |  |  |  |  |  |
| Kalpouzos et al., 2012  [20] | 63 | 2405 | 520 | 1 | = | No | No | 1504 | (24 -90 -42) to (40 -72 -30) centered at (33 -81 -37) |
|  |  |  |  | 2 | ↑ | No | No | 1472 | (28 -70 -34) to (42 -56 -22) centered at (35 -63 -29) |
|  |  |  |  | 3 | ↑ | No | No | 1312 | (-48 -86 -44) to (-28 -68 -34) centered at (-35 -77 -40) |
|  |  |  |  | 4 | = | No | No | 1112 | (-10 -78 -22) to (8 -66 -10) centered at (1 -72 -15) |
|  |  |  |  | 5 | ↓ | No | No | 792 | (-48 -54 -46) to (-34 -36 -36) centered at (-40 -42 -41) |
| Maguire et al., 2003  [21] | 63 | 2417 | 496 | 1 | = | No | No | 1504 | (24 -90 -42) to (40 -72 -30) centered at (33 -81 -37) |
|  |  |  |  | 2 | = | No | No | 1472 | (28 -70 -34) to (42 -56 -22) centered at (35 -63 -29) |
|  |  |  |  | 4 | = | No | No | 1104 | (-42 -86 -44) to (-28 -68 -34) centered at (-33 -77 -40) |
|  |  |  |  | 3 | ↓ | No | No | 1112 | (-10 -78 -22) to (8 -66 -10) centered at (1 -72 -15) |
|  |  |  |  | 5 | = | No | No | 808 | (-48 -54 -46) to (-34 -36 -36) centered at (-40 -42 -41) |
| Salami et al., 2012  [22] | 63 | 2149 | 504 | 1 | = | No | No | 1504 | (24 -90 -42) to (40 -72 -30) centered at (33 -81 -37) |
|  |  |  |  | 5 | ↓ | Yes | No | 624 | (28 -70 -32) to (40 -62 -22) centered at (35 -66 -27) |
|  |  |  |  | 2 | ↓ | No | No | 1312 | (-46 -86 -44) to (-28 -68 -34) centered at (-35 -77 -40) |
|  |  |  |  | 3 | = | No | No | 1112 | (-10 -78,-22) to (8,-66,-10) centered at (1 -72 -15) |
|  |  |  |  | 4 | = | No | No | 808 | (-48 -54 -46) to (-34 -36 -36) centered at (-40 -42 -41) |
| Steffener et al., 2009  [23] | 63 | 2389 |  | 1 | = | No | No | 1504 | (24 -90 -42) to (40 -72 -30) centered at (33 -81 -37) |
|  |  |  |  | 2 | = | No | No | 1472 | (28 -70 -34) to (42 -56 -22) centered at (35 -63 -29) |
|  |  |  |  | 3 | ↑ | No | No | 1312 | (-48 -86 -44) to (-28 -68 -34) centered at (-35 -77 -40) |
|  |  |  |  | 5 | = | No | No | 808 | (-48 -54 -46) to (-34 -36 -36) centered at (-40 -42 -41) |
|  |  |  |  | 4 | ↓ | No | No | 672 | (-10 -76 -22) to (4 -66 -12) centered at (1 -72 -16) |
| Thürling et a., 2014  [24] | 45 | 2407 | 560 | 1 | ↓ | No | No | 1232 | (24 -90 -42) to (40 -76 -30) centered at (33 -83 -37) |
|  |  |  |  | 2 | ↓ | No | No | 848 | (26 -68 -34) to (40 -56 -28) centered at (34 -61 -30) |
|  |  |  |  | 5 | ↓ | Yes | No | 600 | (-38 -86 -42) to (-28 -72 -34) centered at (-33 -81 -39) |
|  |  |  |  | 4 | ↓ | Yes | No | 648 | (-2 -78 -18) to (8 -66 -10) centered at (4 -72 -15) |
|  |  |  |  | 3 | ↓ | No | No | 832 | (-48 -54 -46) to (-34 -36 -36) centered at (-40 -42 -41) |
| Yu et al., 2017  [3] | 55 | 2003 | 584 | 4 | ↓ | Yes | No | 656 | (28 -80 -42) to (40 -72 -36) centered at (34 -76 -39) |
|  |  |  |  | 1 | ↑ | No | No | 1592 | (28 -70 -34) to (42 -56 -22) centered at (35 -63 -29) |
|  |  |  |  | 2 | ↓ | No | No | 920 | (-48 -84 -40) to (-30 -68 -38) centered at (-36 -75 -41) |
|  |  |  |  | 3 | ↓ | No | No | 776 | (-10 -78,-24) to (8,-66,-10) centered at (1 -73 -16) |
| Cluster 5 from main analysis does not survive | | | | |  |  |  |  |  |
| Ziegler et al., 2012  [25] | 59 | 1894 | 488 | 1 | ↑ | No | No | 1544 | (24 -90 -42) to (40 -72 -30) centered at (33 -81 -37) |
|  |  |  |  | 2 | ↑ | No | No | 1544 | (26 -70 -34) to (42 -56 -22) centered at (35 -63 -29) |
|  |  |  |  | 3 | ↑ | No | No | 1352 | (-48 -86 -44) to (-28 -68 -34) centered at (-35 -77 -40) |
|  |  |  |  | 4 | ↑ | No | No | 1328 | (-10 -82,-30) to (8,-66,-10) centered at (0 -73 -17) |
|  |  |  |  | 5 | ↑ | No | No | 824 | (-48 -54 -46) to (-34 -36 -36) centered at (-40 -42 -41) |
| *Alzheimer’s disease* | | | | | | | | | |
| None | 35 | 529 | 440 | 1 | NA | NA | NA | 1144 | (26 -78 -40) to (34 -62 -24) centered at (30 -68 -32) |
|  |  |  |  | 2 | NA | NA | NA | 856 | (30 -66 -32) to (44 -58 -20) centred at (38 -61 -25) |
| Ahmed et al., 2019  [26] | 33 | 513 | 512 | 1 | ↓ | Yes | No | 1056 | (26 -78 -40) to (34 -62 -24) centered at (30 -71 -32) |
|  |  |  |  | 2 | ↓ | No | No | 808 | (30 -66 -32) to (44 -58 -20) centered at (38 -61 -25) |
| Canu et al., 2011  [5] | 31 | 512 | 568 | 1 | ↓ | No | No | 688 | (26 -78 -40) to (34 -68 -24) centered at (30 -73 -29) |
| Canu et al., 2012  [7] | 34 | 505 | 528 | 1 | ↓ | No | No | 808 | (30 -66 -32) to (44 -58 -20) centered at (38 -61 -25) |
| Colloby et al., 2014  [27] | 33 | 482 | 496 | 1 | ↓ | No | No | 1088 | (26 -78 -40) to (34 -62 -24) centered at (30 -71 -32) |
|  |  |  |  | 2 | ↓ | No | No | 840 | (30 -66 -32) to (44 -58 -20) centered at (40 -62 -24) |
| Farrow et al., 2006  [28] | 33 | 522 | 552 | 1 | ↓ | No | No | 1056 | (26 -78 -40) to (34 -62 -24) centered at (30 -71 -32) |
|  |  |  |  | 2 | ↓ | No | No | 808 | (30 -66 -32) to (44 -58 -20) centered at (38 -61 -25) |
| Guo et al., 2016  [6] | 31 | 495 | 544 | 1 | ↓ | No | No | 840 | (30 -66 -32) to (44 -58 -20) centred at (38 -61 -25) |
| Lehmann et al., 2011  [29] | 34 | 499 | 440 | 1 | ↑  (clusters 1 and 2 merged) | Yes | No | 2080 | (26 -78 -40) to (44 -58 -20) centered at (33 -67 -29) |
| Mazere et al., 2007  [30] | 33 | 521 | 504 | 1 | ↓ | No | No | 1056 | (26 -78 -40) to (34 -62 -24) centered at (30 -68 -32) |
|  |  |  |  | 2 | ↓ | Yes | No | 800 | (30 -66 -32) to (44 -58 -20) centered at (38 -61 -25) |
| Möller et al., 2013  [8]^b^ | 27 | 409 | 608 | 1 | ↓ | Yes | No | 624 | (24 -78 -30) to (34 -68 -24) centered at (30 -74 -27) |
| Ossenkoppele et al., 2015  [31] | 33 | 471 | 424 | 1 | ↑ (clusters 1 and 2 merged) | Yes | No | 2104 | (26 -78 -40) to (44 -58 -20) centered at (33 -67 -29) |
| Raji et al., 2009  [32] | 32 | 496 | 440 | 1 | ↑ (clusters 1 and 2 merged) | Yes | No | 2128 | (26 -78 -40) to (44 -58 -20) centered at (33 -67 -29) |
| Serra et al., 2014  [33] | 34 | 481 | 432 | 1 | ↑ (clusters 1 and 2 merged) | Yes | No | 2080 | (26 -78 -40) to (44 -58 -20) centered at (33 -67 -29) |
| Toniolo et al., 2018  [9] | 32 | 442 | 424 | 1 | ↑ (clusters 1 and 2 merged) | Yes | No | 1512 | (26 -78 -40) to (42 -58 -24) centered at (31 -69 -31) |

**^b^In the jackknife analysis for Möller et al. (2013), the large AD cluster split into two subclusters (numbered 1 and 2 here) that were no longer contiguous but remained in the same spatial location as the large combined cluster. Another third cluster in left anterior cerebellar regions emerged as significant which had not been found in any of the other analyses.**

**Table D2. Summary of the effects of jackknifing on each of the clusters from the main ageing and AD analyses.**

| **Cluster from main analysis** | **Survives thresholding in *n* one-study removed analyses**  **(%)** | **Is reduced in size in *n* one-study removed analyses**  **(%)** | **Is increased in size in *n* one-study removed analyses**  **(%)** | **Is unchanged in size in *n* one-study removed analyses**  **(%)** | **Changes peak location by >2 in *n* one-study removed analyses**  **(%)** | **Changes peak label in *n* one-study removed analyses**  **(%)** |
| --- | --- | --- | --- | --- | --- | --- |
| ***Healthy ageing*** |  |  |  |  |  |  |
| 1 | 18  (100) | 7  (39) | 2  (11) | 9  (50) | 1  (6) | 0  (0) |
| 2 | 18  (100) | 6  (33) | 9  (50) | 3  (17) | 1  (6) | 0  (0) |
| 3 | 17  (94) | 5  (28) | 8  (44) | 4  (22) | 3  (17) | 0  (0) |
| 4 | 17  (94) | 5  (28) | 3  (17) | 9  (50) | 0  (0) | 0  (0) |
| 5 | 15^a^  (83) | 4  (22) | 1  (6) | 10  (56) | 1  (6) | 0  (0) |
| ***Alzheimer’s disease*** | | | | | | |
| 1 | 11^b^  (85) | 5  (45) | 6^c^  (55) | 0  (0) | 7  (63)^d^ | 0  (0) |
| 2 | 11^b^  (85) | 5  (45) | 6^c^  (55) | 0  (0) | 6  (55)^d^ | 0  (0) |

^a^This cluster splits into two smaller clusters that are no longer connected in one of the jackknife analyses.

Note that changes in size here refer to any alterations in the cluster size, regardless of whether this change was in one voxel or a large number of voxels.

^b^This includes instances in which cluster 1 and 2 merge, which are counted as survival of each cluster. ^c^This refers to cases in which clusters 1 and 2 merged.

^d^This includes five instances in which clusters 1 and 2 merge.

Percentages of changes in cluster properties are based on all instances in which this cluster survives.

**a.**

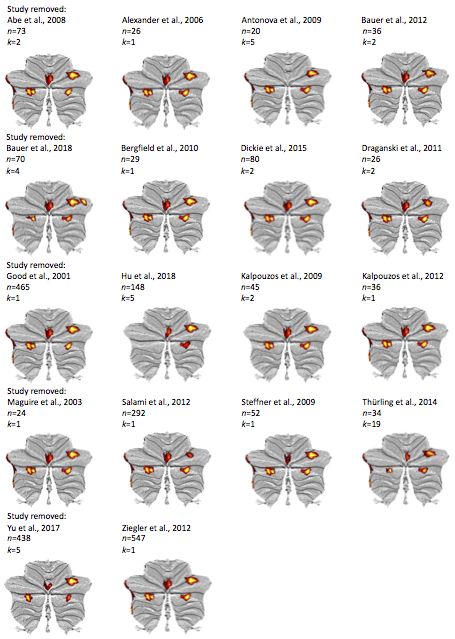

**b.**

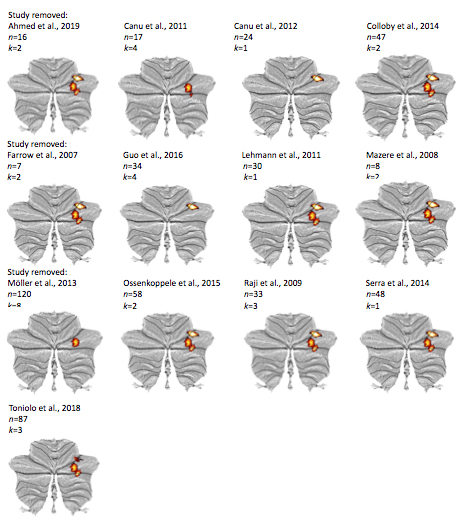

**Figure D1. Flatmaps for each of the 18 jackknife analyses for age-related grey matter decline (a) and for the 13 jackknife analyses for atrophy associated with Alzheimer’s disease (b). The number of subjects included in each study is indicated by *n* and the number of coordinates of cerebellar grey matter loss is given by *k*.**

**a.**

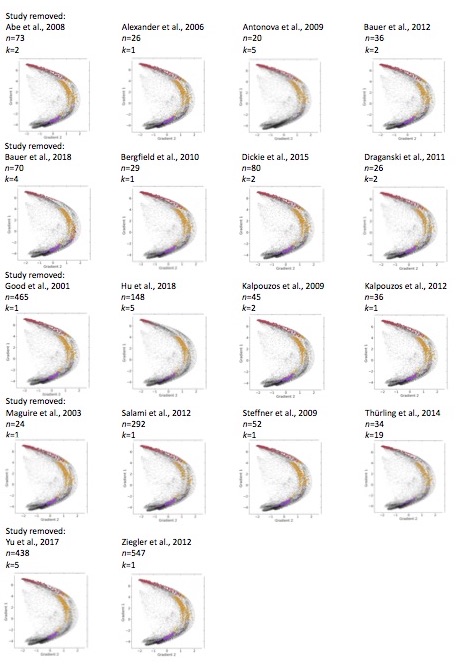

**b.**

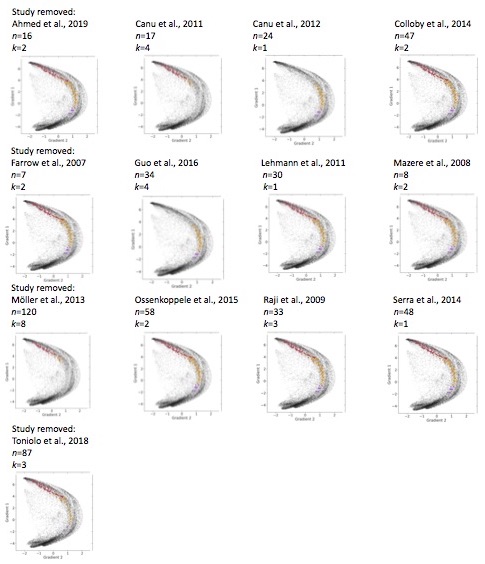

**Figure D2. Gradient 1 and 2 values plotted for each of the 18 jackknife analyses of age-related grey matter decline (a) and the 13 analyses for AD (b). The number of subjects included in each study is indicated by *n* and the number of coordinates of cerebellar grey matter loss is given by *k*.**

**a.**

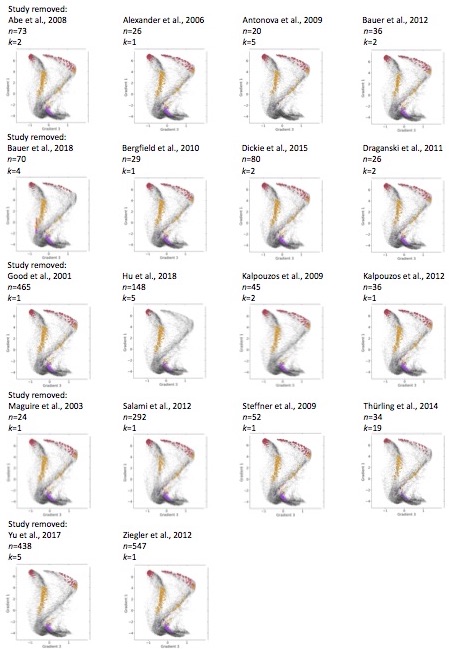

**b.**

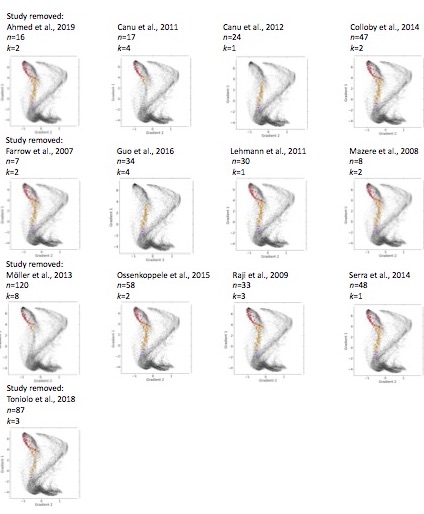

**Figure D3. Gradient 1 and 3 values plotted for each of the 18 jackknife analyses of age-related grey matter decline (a) and the 13 jackknife analyses for AD (b). The number of subjects included in each study is indicated by *n* and the number of coordinates of cerebellar grey matter loss is given by *k.***

**13 Alzheimer’s disease jackknife tests	18 healthy ageing jackknife tests**

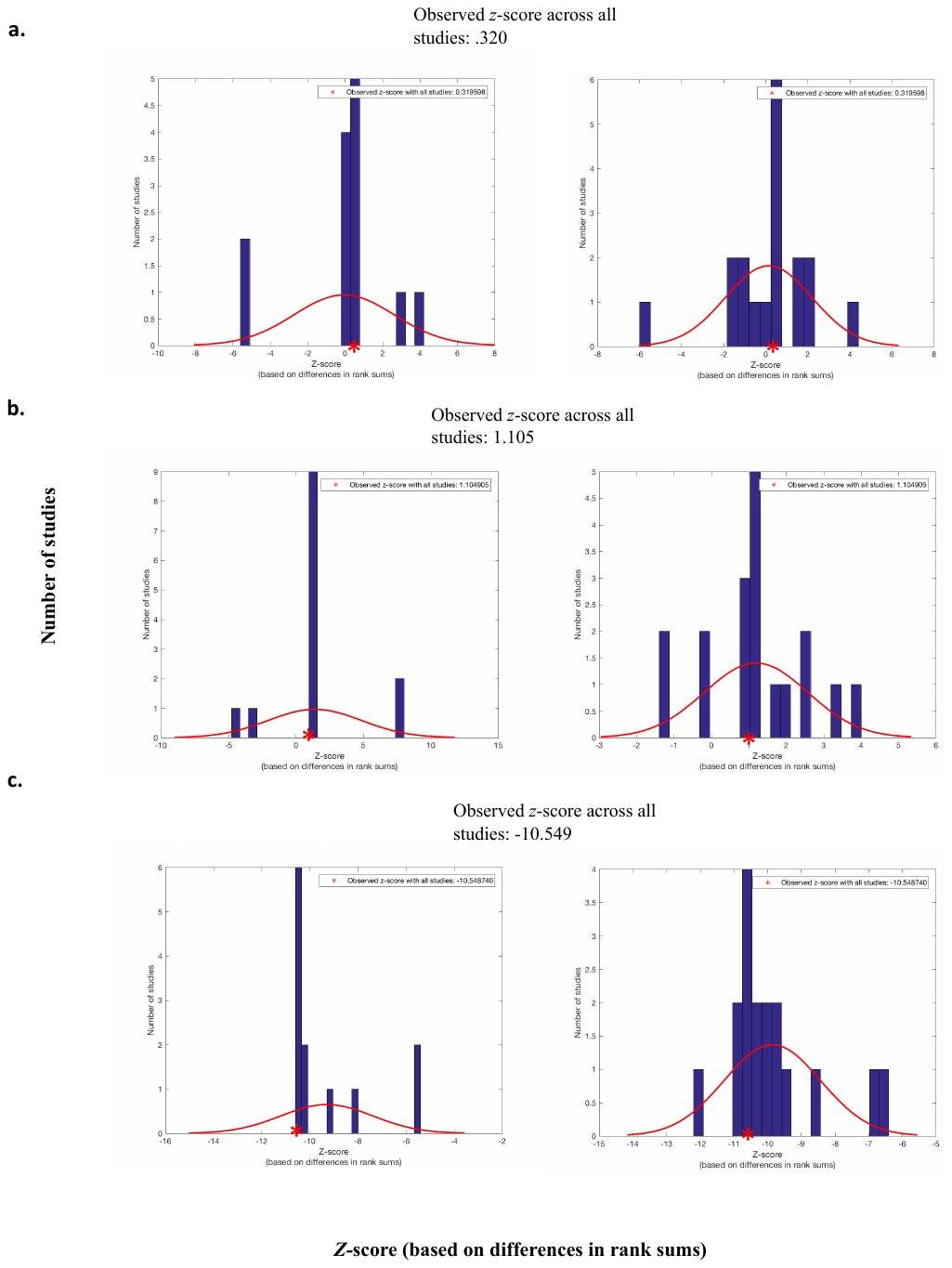

**Figure D4. Summary of differences in gradient values between AD and healthy ageing following a jackknife procedure.**

**Panels a, b, and c represent jackknife analyses for gradients 1, 2, and 3, respectively. The differences between gradient values for AD and ageing were calculated as *z-*scores based on the Wilcoxon rank sum test. Left panels show results of comparing the gradient values from the total healthy ageing sample (18 studies) to the gradient values from the Alzheimer’s disease data set with a given study removed from the analysis (i.e. 12 studies rather than the full 13). Right panels show results of comparing the gradient values from the total AD sample (13 studies) to the gradient values from the healthy ageing data set with a given study removed from the analysis (i.e. 17 studies rather than the full 18). The red asterisk marks the observed mean difference between all AD and all healthy ageing studies.**
